# Changes in affect variability after starting gender-affirming hormone therapy

**DOI:** 10.1101/2024.08.28.24312697

**Authors:** Margot W. L. Morssinkhof, Marijn Schipper, Baudewijntje P. C. Kreukels, Karin van der Tuuk, Martin den Heijer, Odile A. van den Heuvel, David Matthew Doyle, Birit F. P. Broekman

## Abstract

Affect variability is determined by how often and how strongly negative affect changes over time. Cisgender women report greater variability in affect than cisgender men. It has been suggested that sex hormone changes may influence affect variability. Transgender people frequently opt to use sex hormones in the form of gender-affirming hormone therapy (GAHT), but the extent to which GAHT can change negative affect variability is not yet clear. Therefore, this study aims to study changes in negative affect variability after starting GAHT.

We have included data from 94 participants from the RESTED study: 49 transmasculine (TM) participants (assigned female at birth, starting testosterone) and 45 transfeminine (TF) participants (assigned male at birth, starting estrogens and anti-androgens). Participants completed up to 7 consecutive daily diaries at each of three time points: before starting GAHT, and after 3 and 12 months of GAHT. The daily diaries collected participants’ reports on symptoms related to negative affect: experienced low mood, less interest, tense feelings and restless feelings. We have used linear mixed models to compare negative affect variability during one week, corrected for mean negative affect, between gender groups (TM versus TF) and measurement time points.

Results show that in the TM group, variability in low mood, tense feelings and restless feelings decreases after 12 months of GAHT. In the TF group, variability in low mood increases after 3 months and 12 months of GAHT, as does variability in restless feelings after 3 months of GAHT. Group comparisons indicate significant group differences in changes in variability in low mood and restless feelings, with stronger increases in variability of negative affect in the TF group compared to TM group after 3 and 12 months of GAHT.

Our findings indicate that variability patterns in negative affect in transgender persons tend to cross- over from being consistent with sex assigned at birth before GAHT to being more in line with gender identity after 12 months of GAHT. Future studies should focus on measuring both negative and positive affect variability during GAHT, preferably through multiple measurements per day, taking into account diverse social and daily contextual factors during GAHT.

**Graphical abstract:** 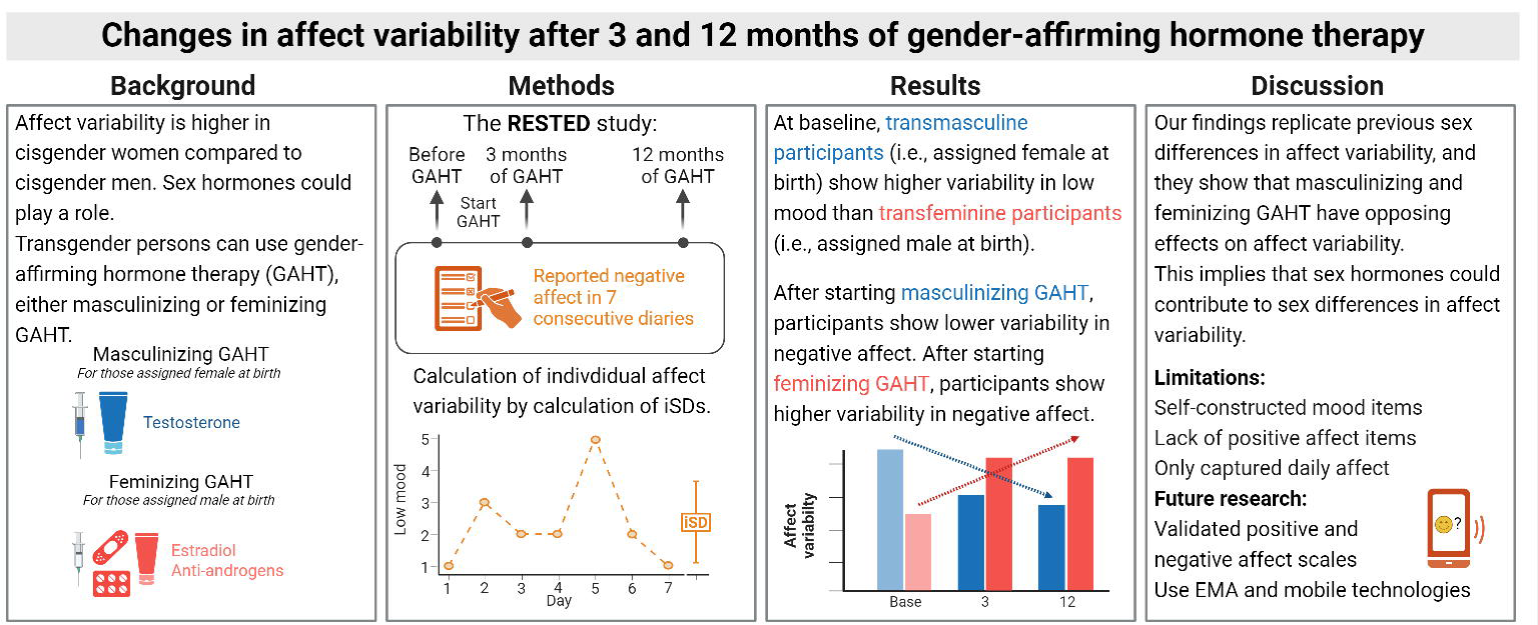

**Highlights:**

◦ Cisgender women report greater affect variability than cisgender men.
◦ The impact of GAHT on affect variability in transgender people is unclear.
◦ We examined affect variability in transmasculine (TM) and transfeminine (TF) people.
◦ We find decreased variability in negative affect after starting masculinizing GAHT.
◦ We find increased variability in negative affect after starting feminizing GAHT.

## Introduction

Emotions color our everyday lives. The emotional landscape is shaped by both mood and affect, which can be conceptually distinguished from one another. Mood represents the enduring emotional atmosphere (‘’climate’’), while affect captures the transient emotions we experience moment-to- moment (‘’weather’’) (Manjunatha et al., 2009). Affect intensity is determined by how strongly we feel a specific emotion at a given time, while affect variability is determined by how often and how strongly affect changes over time (Larsen & Diener, 1987; Maciejewski et al., 2015). Variability in positive and negative affect seems to be somewhat trait-like, since individuals who report stronger variability in negative moods also tend to report stronger variability in positive moods (Larsen & Diener, 1987). Studies investigating emotional changes often focus on assessing overall mood, but measuring affect variability can provide deeper insights into our daily emotional well-being (Wang et al., 2012).

Studies find sex and/or gender differences in variability in affect, with cisgender women reporting more affect variability compared to cisgender men (Neiss & Almeida, 2004; Wang et al., 2012). There could be numerous reasons for this difference. First, this could be due to gender differences in self-reported emotions, which can be affected by gendered beliefs about emotion, internalized gender stereotypes, and social positions (Robinson & Clore, 2002) or by better emotional awareness in women compared to men (Mankus et al., 2016).

Second, differences in affect variability could be related to biological differences between cisgender women and men. This is seen in the menstrual cycle, in which women with premenstrual syndrome experience increased variability in both positive and negative affect in the premenstrual week (Bowen et al., 2011). Sex hormone interventions can also change affect: suppression of sex hormone production results in increased affect variability (defined in the article as mood lability measured with the Profile of Mood States or POMS) in healthy women in an experimental setting (Stenbæk et al., 2016) and in men with prostate cancer undergoing androgen deprivation therapy (Donovan et al., 2015). Women who use oral contraceptives, which contain synthetic progestins and estrogens, show lower affect variability than naturally cycling women, which has been called “emotional blunting” (Hamstra et al., 2017). Other studies, however, did not replicate this (Weigard et al., 2021).

Research on the effects of sex hormones on affect variability is still inconsistent, but possible effects of sex hormone changes are relevant for transgender people who start gender-affirming hormone therapy (GAHT). Transgender people experience a discrepancy between their gender identity and their sex assigned at birth. They can use GAHT to better align their physical characteristics with their gender identity: those assigned female at birth with a masculine gender identity can use masculinizing GAHT, which consists of testosterone, and those assigned male at birth with a feminine gender identity can use feminizing GAHT, which consists of estrogens and anti-androgens.

Numerous studies indicate that use of GAHT could change affect variability (Doyle et al., 2023). Qualitative work by Wassersug et al. (2007) found affect changes after use of feminizing GAHT, with a participant describing her experience as “an emotional rollercoaster ride”. Van Goozen et al. (1995) is the only quantitative study examining the effects of GAHT using diary-style reported affect outcomes. Participants completed daily questionnaires for 15 days, each containing 15 items with a 10-point intensity rating scale. The authors found that after 3 months of GAHT, transfeminine (TF) individuals rated the item “changeable mood” higher than transmasculine (TM) individuals. However, the authors report that they averaged reported so-called “changeable mood” over all 15 days, meaning insights in day-to-day affect variability is still missing.

In studies on affect variability, it is important to distinguish between different methods used to measure affect variability. These methods differ both in how affect variability is reported and in which timeframe this happens. Affect variability can be measured through questionnaires or questions which directly require participants to report their fluctuations in affect. For example, the Affective Lability Scale (Harvey et al., 1989) includes items like “One minute I can be feeling OK and then the next minute I’m tense, jittery, and nervous”. However, asking participants to retrospectively estimate these outcomes means that reports are most likely based on heuristic estimation processes (Solhan et al., 2009), which can be affected by one’s self image and internalized stereotypes or judgements (Robinson & Clore, 2002).

A second method is to ask participants about their momentary affect levels (i.e., “How tense do you feel?”) at multiple points over time. Affect variability can then be calculated based on the differences in momentary affect levels over time. Using this method, studies differ in the timeframe in which this affect is reported or recalled. Some studies ask participants to estimate their affect retrospectively over set timespan (i.e., “How sad have you felt this week/today/in the past hours?”), whereas others ask “online” affect levels (i.e. “How sad do you feel right now?”) (Trull & Ebner-Priemer, 2013).

Retrospectively reported affect variability, as seen in Van Goozen et al. (1995), is only modestly associated with affect variability as measured using momentary affect measurements (Solhan et al., 2009).

In this study, we prospectively investigate changes in negative affect variability at 3 and 12 months after starting masculinizing or feminizing GAHT in transgender persons. We firstly examine whether TM participants and TF participants report differences in negative affect variability before starting GAHT. Secondly, we examine whether affect variability changes after 3 and 12 months of masculinizing and feminizing GAHT, and thirdly, we examine whether changes after GAHT differ between masculinizing and feminizing GAHT users. We hypothesize that TM participants report higher negative affect variability than TF participants at baseline. We further hypothesize that changes in affect variability will differ in those starting masculinizing or feminizing hormones, with a decrease in negative affect variability in participants starting masculinizing hormones and an increase in negative affect variability in participants starting feminizing hormones.

## Methods

### Participants

For this study, we include data from participants who took part in the RESTED study (Morssinkhof et al., 2023), a prospective study which aimed to investigate changes in sleep and mood after 3 and 12 months of GAHT use. All participants were recruited at the Amsterdam University Medical Centre (AUMC) or University Medical Centre Groningen (UMCG), were proficient in the Dutch language, did not use any sleeping medication, and were aged between 18 and 50 years old. The upper age limit was included in the inclusion criteria since the sleep measurement device used in the study was validated up to the age of 50: for more information, see Morssinkhof et al. (2023). The study was approved by the medical ethical committee of the Amsterdam UMC (location VUmc), which declared that the Medical Research Involving Human Subjects Act (WMO) did not apply (study number 2019.353). Study data was collected between January 2020 and September 2023.

Detailed descriptions of study recruitment, data collection and study outcomes for the RESTED study can be found in Morssinkhof et al. (2023) and Morssinkhof et al. (2024). For the current study, we use data from daily diaries on negative affect from 7 consecutive days collected at each measurement time point (i.e., at baseline, after 3 months of GAHT and after 12 months of GAHT). To ensure that we have adequate samples for the within-person calculation of affect variability (Maciejewski et al., 2015), we exclude measurements in which participants completed fewer than 3 daily diaries. This results in exclusion of 5 participants. Figure 1 displays the full participant flowchart, including sample sizes at baseline and follow-up.

**Figure 1.**
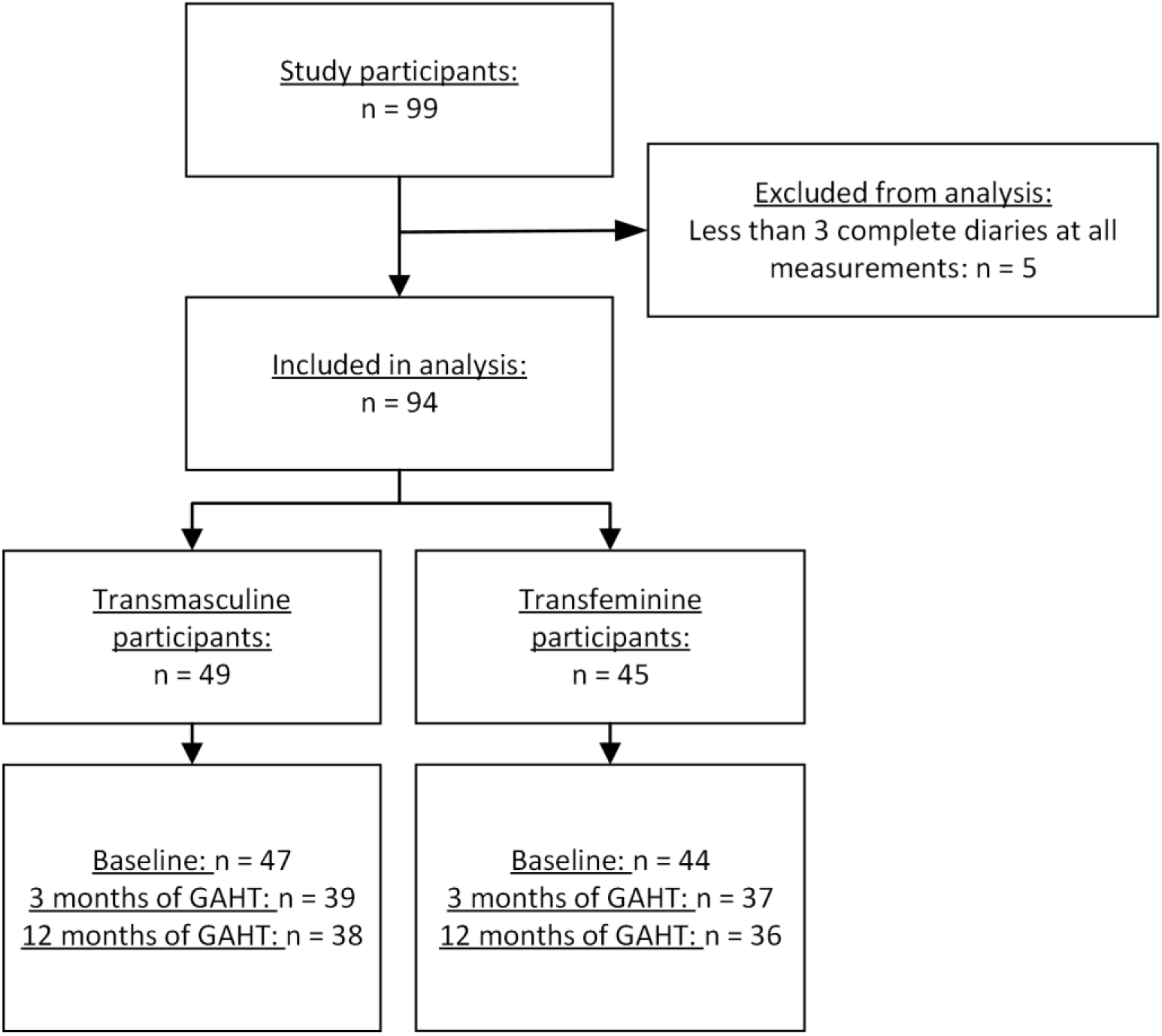
Flowchart displaying sample sizes at inclusion, in the study database and at each follow-up moment. A total of 39 TM participants contributed a baseline measurement and at least one follow-up measurement, and a total of 40 TF participants contributed a baseline measurement and at least one follow-up measurement.

### Gender-affirming hormone therapy

Participants received GAHT in accordance with the Standards of Care of the World Professional Association for Transgender Health (WPATH) (Coleman et al., 2022). All participants started GAHT after the baseline measurement of the study. TM participants, who were assigned female at birth, started using testosterone, either in transdermal form (Androgel, daily dose of 40.5mg) or intramuscular form (testosterone esters, 250 mg every 3 weeks; testosterone undecanoate, 1000 mg every 12 weeks). Some TM participants also started or continued use of cycle regulation medication or hormonal contraceptives, consisting of progestins (lynestrenol, 5 mg; desogestrel, 0.075mg; levonorgestrel IUD, 52mg; medroxyprogesterone, 150 mg) or combined oral contraceptives (ethinylestradiol/levonorgestrel, 0.02/0.10 mg; ethinylestradiol/levonorgestrel, 0.03/0.15 mg; ethinylestradiol/drospirenone 0.02/3 mg). Most TF participants, who were assigned male at birth, started using estrogens, either in transdermal (estradiol gel, 0.06% 1.5 mg daily; or estradiol patches, 100 mcg per 24 hours, twice a week) or oral (estradiol valerate, 2mg twice daily) form, as well as anti-androgens, either in the form of oral cyproterone acetate (daily dose of 10 mg) or GnRH-analogues (triptorelin 3.75 mg per 4 weeks; leuproreline, 3.75 mg per 4 weeks; triptorelin 11.25 mg per 12 weeks). Three TF participants started using only estrogens (n = 1) or only anti-androgens (n = 2); we have conducted a sensitivity analysis excluding these participants.

As part of clinical care, serum testosterone and estradiol levels were measured prior to start of treatment as well as after 3 and 12 months of GAHT. In both participating centers, serum hormone levels were analyzed using liquid chromatography-tandem mass spectrometry (LC-MS/MS). The LC-MS/MS assays for testosterone had a lower limit of quantitation of 0.1 nmol/L, and an inter-assay coefficient of variation of 4% to 9%. The LC-MS/MS assays for estradiol had a lower limit of quantitation of 20 pmol/L and an inter-assay coefficient of variation of <7%.

### Demographic and outcome variables

#### Baseline characteristics

We have obtained participants’ sex assigned at birth, gender identity, age, alcohol consumption, smoking status, psychotropic medication use and the form and dosage of GAHT use from institutional medical records. We have further categorized psychotropic medication into antidepressants, anxiolytics, antipsychotics and/or stimulants, and cycle regulation into progestin-only forms (i.e., levonorgestrel intrauterine device, medroxyprogesterone injections, desogestrel implant, oral progestins) and combined forms (i.e., combined oral contraceptives containing estrogens and progestins).

#### Outcome variables

Participants received 7 consecutive daily diaries at each time point of the study (i.e., before starting GAHT, after 3 months of GAHT and after 12 months of GAHT). These diaries were sent via email every morning and comprised a total of 32 items. The items were part of the original Consensus Sleep Diary covering sleep, mood, and lifestyle (Carney et al., 2012), augmented with additional items, including four self-formulated items of negative affect. The negative affect items were self-constructed and based on key symptoms of depressive and anxiety disorders from the Diagnostic and Statistical Manual of Mental Disorders fifth edition (DSM-5). Depressive symptom items include “Yesterday, I had somber feelings” (Low Mood) and “Yesterday, I had less interest in things I am usually enthusiastic about” (Less interest). Anxiety symptom items include “Yesterday, I had tense feelings” (Tense Feelings) and “Yesterday, I had a restless feeling in my body” (Restless Feeling). All items were rated from 1 to 5 on a Likert scale (1 = “Not at all”; 5 = “Very much”).

For this study, we have calculated negative affect variability and intensity per item, creating within-subject standard deviations (iSD) and within-subject means (iMean) of each item for every measurement time point (i.e., baseline, 3 months of GAHT and 12 months of GAHT).

The iSD alone does not account for chronological order of daily mood reports, meaning it cannot capture distinctions between frequent changes in negative affect (e.g., scores of 5, 2, 4, 2, 5) and gradual negative affect changes (e.g., scores of 5, 5, 4, 2, 2). We have therefore additionally calculated the Mean Absolute Successive Differences (MASDs) for each participant at every measurement time point, which are shown in the Supplementary materials 1 (Supplementary Table 2). The MASD can account for affect changes over consecutive days, serving as a complementary measure for within-participant negative affect variability (Maciejewski et al., 2015). Figure 2 displays an illustration of the calculation of the iMean, iSD and MASD for two of the study participants.

**Figure 2.**
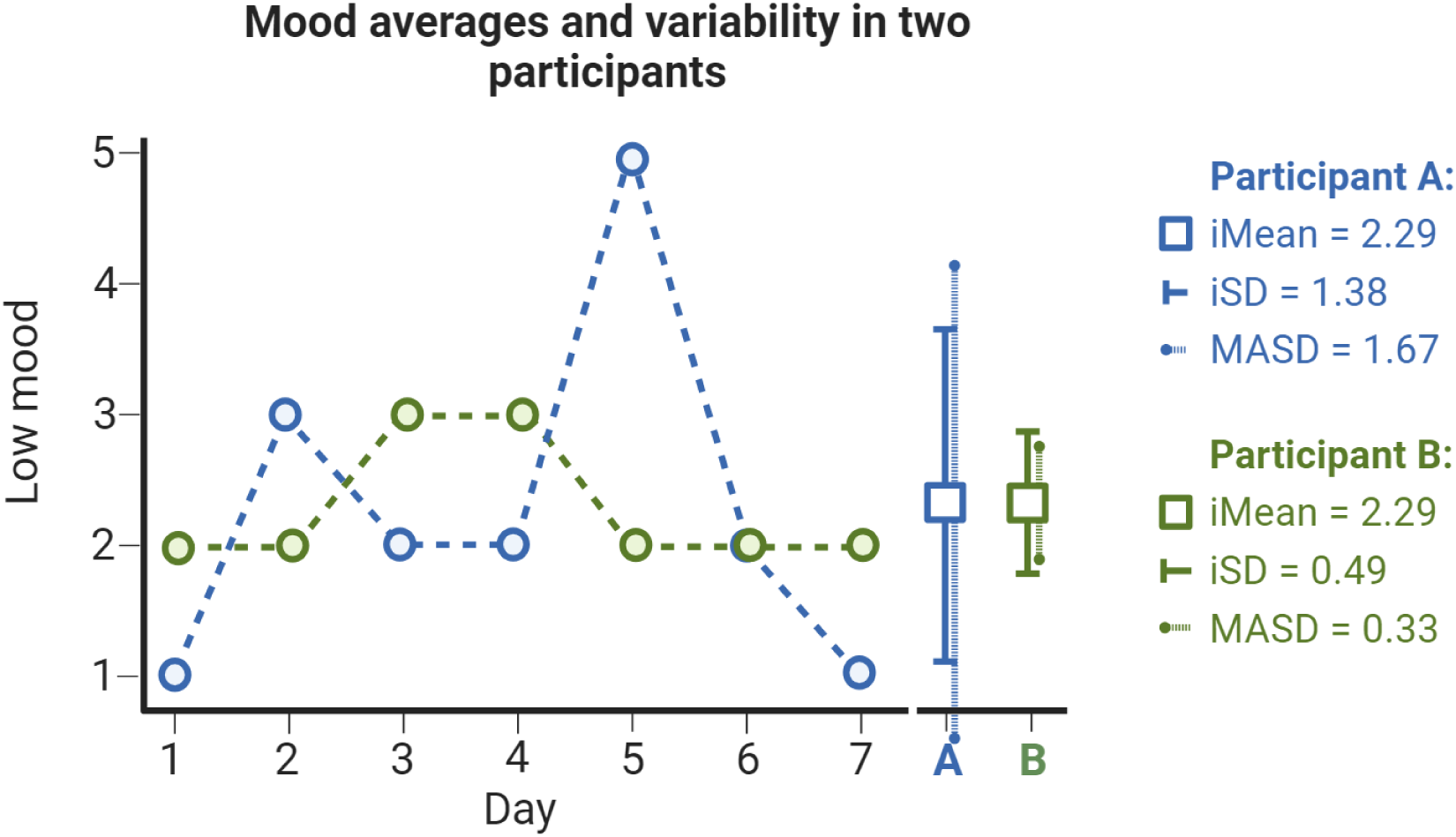
Illustration of the calculations of negative affect scores, averages and variability. The iMean is the within-person mean level of the low mood reports in the 7 diaries, the iSD is the within-person SD of the low mood reports in the 7 diaries and the MASD is the Mean Absolute Successive Difference of the low mood reports in the 7 diaries, which is calculated by the changes in day-to-day levels of low mood. The figure displays two participants in the database who report the same average for negative affect, but participant A has more variability in negative affect compared to participant B.

Since affect variability is based on the deviation around the mean, this outcome is particularly sensitive to floor or ceiling effects, especially in participants who experience low levels of negative affect (von Klipstein et al., 2023). Therefore, we have corrected all models for average negative affect.

### Statistical analyses

All statistical analyses have been conducted in Rstudio (R Development Core Team, 2010), using the lme4 (Bates et al., 2009) and lmerTest packages (Kuznetsova et al., 2017).

To analyze differences in negative affect variability between TM and TF participants before starting GAHT, we use linear regression analyses. These analyses include the iSD at baseline as the outcome variable, the group (i.e. feminizing or masculinizing GAHT) as fixed predictor and the iMean as a covariate. Second, to investigate changes in negative affect variability, we analyze TM and TF groups separately, using the iSD as the outcome variable, the measurement time point as categorical predictor and the iMean as a covariate. To analyze potential differences in changes in negative affect variability in both groups, we have added an interaction term for group to the second model. We have run similar models for the iMean and MASD, as reported in Supplementary materials 1.

#### Covariates and sensitivity analyses

We have conducted a sensitivity analysis excluding participants who started using only estrogens (n = 1) or only anti-androgens (n = 2). We have also performed sensitivity analyses excluding participants who had only contributed a baseline measurement or who did not contribute a follow-up measurement (i.e., complete case analysis). To account for possible effects of covariates, we have added a categorical predictor for cycle regulation (i.e., cycle regulation use compared to no cycle regulation use) and psychotropic medication use (i.e. psychotropic medication use compared to no psychotropic medication use) to our analyses.

### Transparency and Openness

Since data of this study contains patient data from a particularly vulnerable group (i.e., transgender participants), anonymized study data is available upon reasonable request by contacting the corresponding author. The study codebook and analysis code for all analyses is available via osf.io/8dnjp/.

## Results

### Demographic and clinical characteristics

We have conducted study analyses on data of 94 participants, who are on average 25 years old (SD 6). A total of 18% use psychotropic medication, and among TM participants 45% use cycle regulation at baseline. Table 1 displays clinical and demographic baseline characteristics of the group.

**Table 1.** Clinical and demographic baseline characteristics of study participants.

|  |  |  |  |
| --- | --- | --- | --- |
|  |  | **Transmasculine group** | **Transfeminine group** |
| **Sample size in total cohort** |  | 49 | 45 |
| **Age** | *Median (IQR)* | 22 (20 to 24) | 26 (23 to 30) |
| **Gender identity** | *Binary\** | 38 (78%) | 43 (96%) |
|  | *Non-binary\** | 11 (22%) | 2 (4%) |
| **Alcohol (consumptions per week)** | *Median (IQR)* | 0 (0 to 1) | 0.5 (0 to 2) |
| **Smoking (n, %)** | *Never* | 40 (82%) | 37 (82%) |
|  | *Previous* | 3 (6%) | 2 (4%) |
|  | *Current* | 5 (10%) | 6 (13%) |
|  | *Missing* | 1 (2%) | 0 (0%) |
| **Psychotropic medication use at baseline (n, %)** | *Antidepressants* | 4 (8%) | 2 (4%) |
|  | *Anxiolytics* | 0 (0%) | 0 (0%) |
|  | *Antipsychotics* | 2 (4%) | 2 (4%) |
|  | *Stimulants* | 3 (6%) | 4 (9%) |
| **Cycle regulation use at baseline (n, %)** | *Progestin-only formulations* | 14 (29%) | - |
|  | *Combined oral contraceptives* | 8 (16%) | - |
| **Form of testosterone prescribed at start of GAHT (n, %)** | *Testosterone gel* | 43 (88%) | - |
|  | *Short-acting injections* | 6 (12%) | - |
|  | *Long-acting injections* | 0 (0%) | - |
| **Form of estrogen prescribed at start of GAHT (n, %)** | *Estradiol Tablets* | - | 27 (60%) |
|  | *Estradiol gel* | - | 4 (10%) |
|  | *Estradiol spray* | - | 0 (0%) |
|  | *Estradiol patches* | - | 13 (29%) |
|  | *None\*\** | - | 1 (2%) |
| **Form of anti-androgens prescribed at start of GAHT (n, %)** | *Cyproterone acetate* | - | 4 (9%) |
|  | *GnRH analogues* | - | 39 (87%) |
|  | *None\*\** | - | 2 (4%) |
| **Testosterone level at start of GAHT\*\*\*** | *N* = 43 | 1.06 nmol/L (SD 1.0) | 15.2 nmol/L (SD 6.0) |
| **Testosterone level after 3 months of GAHT\*\*\*** | *N* = 37 | 24.5 nmol/L (SD 19.5) | 0.94 nmol/L (SD 1.9) |
| **Testosterone level after 12 months of GAHT\*\*\*** | *N* = 36 | 17.6 nmol/L (SD 12.0) | 1.0 nmol/L (SD 1.3) |
| **Estradiol level at start of GAHT\*\*\*** | *N* = 43 | 183.1 pmol/L (SD 182.4) | 75.8 pmol/L (SD 22.2) |
| **Estradiol level after 3 months of GAHT\*\*\*** | *N* = 35 | 170.4 pmol/L (SD 109.1) | 307.9 pmol/L (SD 181.3) |
| **Estradiol level after 12 months of GAHT\*\*\*** | *N* = 35 | 148.4 pmol/L (SD 95.1) | 345.0 pmol/L (SD 201.5) |
| * Binary gender identity: identification as (transgender) man or woman. All participants who did not identify with these gender identities were grouped under non-binary gender identity.<br>** Three TF participants started using only estrogens or anti-androgens, but not both.<br>***For both estrogen and testosterone it must be noted that these can also be affected by use of hormonal contraceptives or progestins. |  |  |  |

### Negative affect variability

Group differences in negative affect variability at baseline are displayed in Table 2, and changes in negative affect variability after GAHT use in Table 3. Figure 3 provides a graphic overview of affect variability (iSD) for each group and time point. The full results on the iSD are reported in this section, and full results on the iMean and MASD are reported in Supplementary Tables 1 and 2.

**Figure 3.**
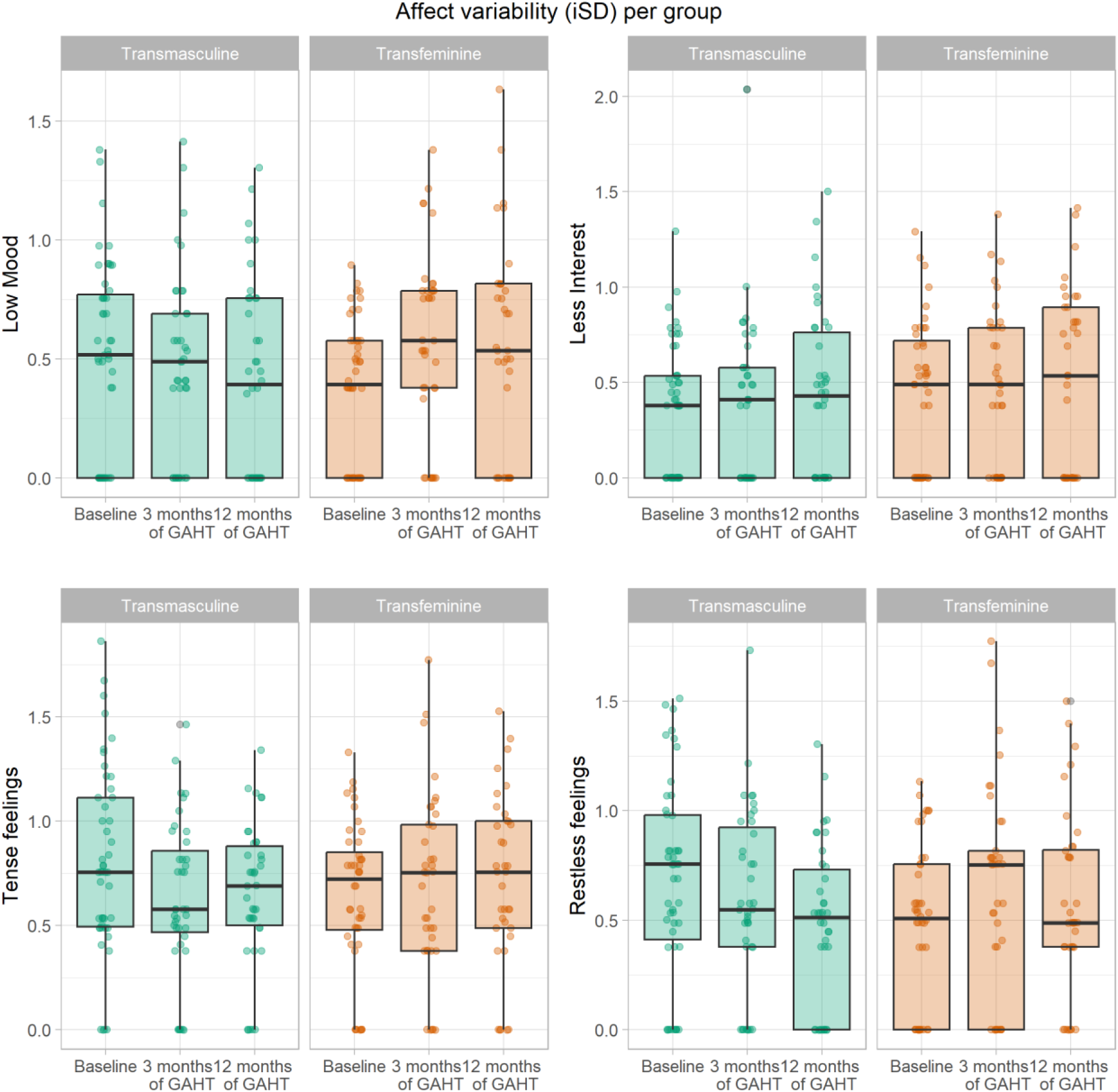
Negative affect variability in the transmasculine and transfeminine group per item at baseline, after 3 months of GAHT and after 12 months of GAHT. The spread of the individual affect variability outcomes is displayed by the dots, and the medians, interquartile ranges and total score ranges are displayed by the boxplots.

**Table 2.** Negative affect variability outcomes before GAHT use (baseline).

|  |  |  |  |  |
| --- | --- | --- | --- | --- |
|  |  | *Transmasculine group* | *Transfeminine group* | *Transfeminine group compared to transmasculine group* |
|  |  | Mean (SD) | Mean (SD) | Estimate, (95% CI), p |
| **Low mood** | iSD | 0.5 (0.40) | 0.37 (0.30) | -0.13<br>**(-0.25 to -0.01)**<br>**P = 0.03** |
| **Less interest** | iSD | 0.37 (0.34) | 0.43 (0.39) | 0.02 (-0.09 to 0.13)<br>P = 0.73 |
| **Tense feelings** | iSD | 0.81 (0.44) | 0.64 (0.36) | -0.13 (-0.29 to 0.03)<br>P = 0.10 |
| **Restless feelings** | iSD | 0.68 (0.45) | 0.46 (0.37) | -0.10 (-0.24 to 0.04)<br>P = 0.16 |
| The transmasculine group is used as the reference group. The iSD estimates are adjusted for the iMeans per item. |  |  |  |  |

**Table 3.**
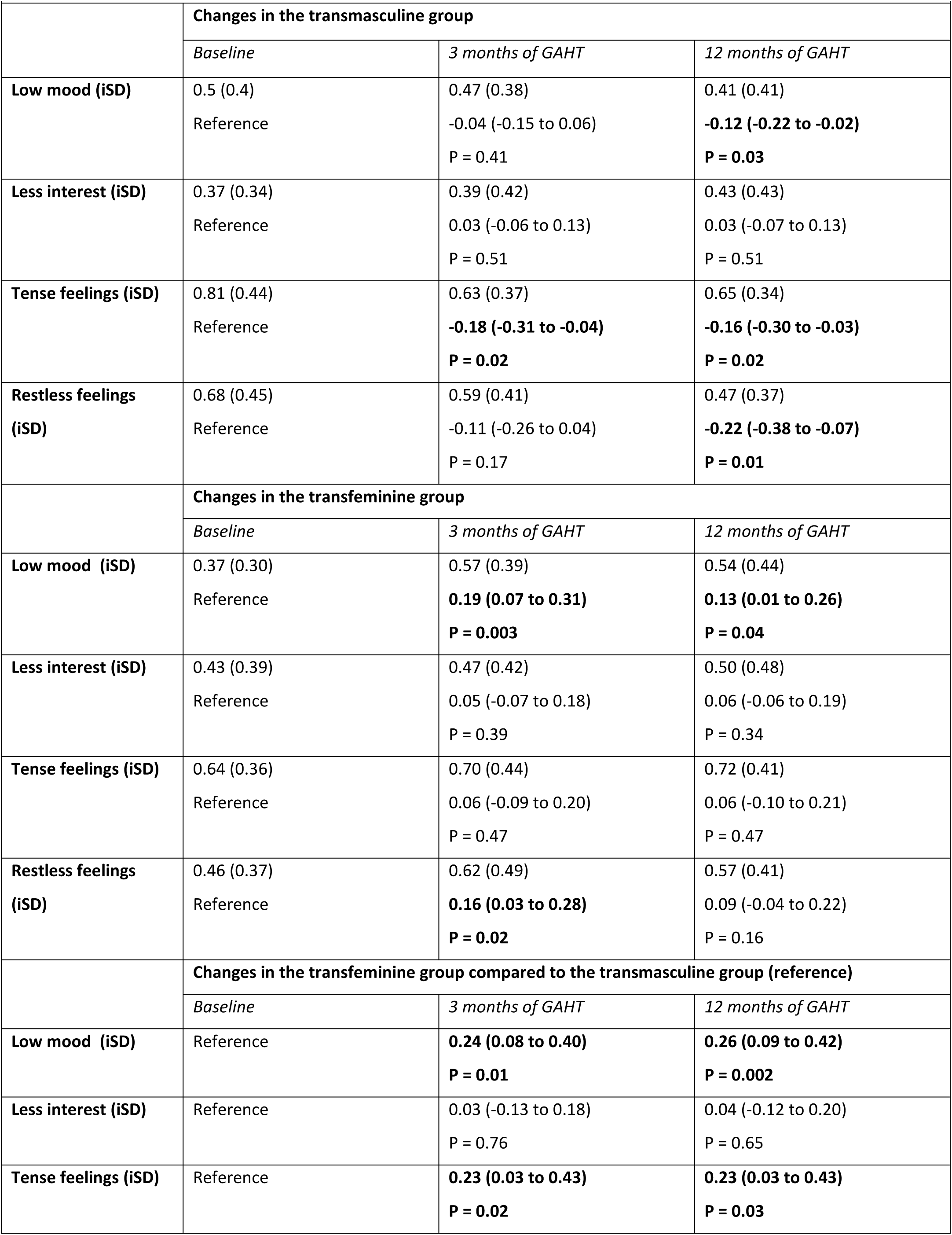
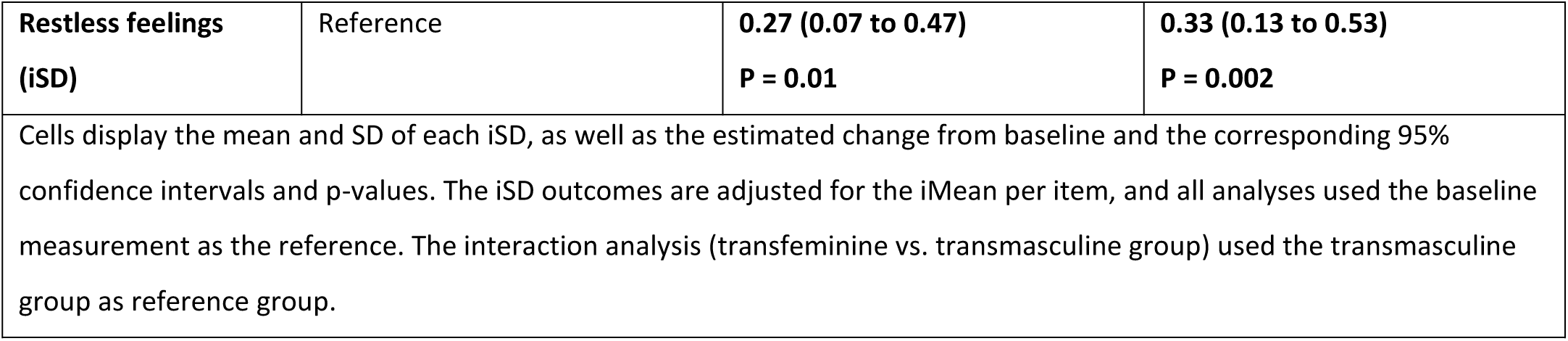
Changes in negative affect variability after GAHT use, stratified per group and with an interaction term for group.

|  |  |  |  |
| --- | --- | --- | --- |
|  | **Changes in the transmasculine group** |  |  |
|  | *Baseline* | *3 months of GAHT* | *12 months of GAHT* |
| **Low mood (iSD)** | 0.5 (0.4)<br>Reference | 0.47 (0.38)<br>-0.04 (-0.15 to 0.06)<br>P = 0.41 | 0.41 (0.41)<br>**-0.12 (-0.22 to -0.02)**<br>**P = 0.03** |
| **Less interest (iSD)** | 0.37 (0.34)<br>Reference | 0.39 (0.42)<br>0.03 (-0.06 to 0.13)<br>P = 0.51 | 0.43 (0.43)<br>0.03 (-0.07 to 0.13)<br>P = 0.51 |
| **Tense feelings (iSD)** | 0.81 (0.44)<br>Reference | 0.63 (0.37)<br>**-0.18 (-0.31 to -0.04)**<br>**P = 0.02** | 0.65 (0.34)<br>**-0.16 (-0.30 to -0.03)**<br>**P = 0.02** |
| **Restless feelings (iSD)** | 0.68 (0.45)<br>Reference | 0.59 (0.41)<br>-0.11 (-0.26 to 0.04)<br>P = 0.17 | 0.47 (0.37)<br>**-0.22 (-0.38 to -0.07)**<br>**P = 0.01** |
|  | **Changes in the transfeminine group** |  |  |
|  | *Baseline* | *3 months of GAHT* | *12 months of GAHT* |
| **Low mood (iSD)** | 0.37 (0.30)<br>Reference | 0.57 (0.39)<br>**0.19 (0.07 to 0.31)**<br>**P = 0.003** | 0.54 (0.44)<br>**0.13 (0.01 to 0.26)**<br>**P = 0.04** |
| **Less interest (iSD)** | 0.43 (0.39)<br>Reference | 0.47 (0.42)<br>0.05 (-0.07 to 0.18)<br>P = 0.39 | 0.50 (0.48)<br>0.06 (-0.06 to 0.19)<br>P = 0.34 |
| **Tense feelings (iSD)** | 0.64 (0.36)<br>Reference | 0.70 (0.44)<br>0.06 (-0.09 to 0.20)<br>P = 0.47 | 0.72 (0.41)<br>0.06 (-0.10 to 0.21)<br>P = 0.47 |
| **Restless feelings (iSD)** | 0.46 (0.37)<br>Reference | 0.62 (0.49)<br>**0.16 (0.03 to 0.28)**<br>**P = 0.02** | 0.57 (0.41)<br>0.09 (-0.04 to 0.22)<br>P = 0.16 |
|  | **Changes in the transfeminine group compared to the transmasculine group (reference)** |  |  |
|  | *Baseline* | *3 months of GAHT* | *12 months of GAHT* |
| **Low mood (iSD)** | Reference | **0.24 (0.08 to 0.40)**<br>**P = 0.01** | **0.26 (0.09 to 0.42)**<br>**P = 0.002** |
| **Less interest (iSD)** | Reference | 0.03 (-0.13 to 0.18)<br>P = 0.76 | 0.04 (-0.12 to 0.20)<br>P = 0.65 |
| **Tense feelings (iSD)** | Reference | **0.23 (0.03 to 0.43)**<br>**P = 0.02** | **0.23 (0.03 to 0.43)**<br>**P = 0.03** |
| **Restless feelings**<br>**(iSD)** | Reference | **0.27 (0.07 to 0.47)**<br>**P = 0.01** | **0.33 (0.13 to 0.53)**<br>**P = 0.002** |
| Cells display the mean and SD of each iSD, as well as the estimated change from baseline and the corresponding 95% confidence intervals and p-values. The iSD outcomes are adjusted for the iMean per item, and all analyses used the baseline measurement as the reference. The interaction analysis (transfeminine vs. transmasculine group) used the transmasculine group as reference group. |  |  |  |

### Negative affect variability at baseline

As shown in Table 2, at baseline the TF group report significantly lower variability for low mood compared to the TM group (-0.13, 95% CI: -0.25 to -0.01, p = 0.03), but other items show no significant group differences.

### Changes in negative affect variability after starting GAHT

Stratified linear mixed models in the TM group indicate that variability in tense feelings decreases after 3 and 12 months of masculinizing GAHT compared to baseline (-0.18, 95% CI: -0.31 to -0.04, p = 0.02; -0.16, 95% CI: -0.30 to -0.03, p = 0.02) and variability in low mood (-0.12, 95% CI: -0.22 to -0.02, p = 0.03) and restless feelings (-0.22, 95% CI: -0.38 to -0.07, p = 0.01) decreases after 12 months of GAHT compared to baseline. Stratified analyses in the TF group shows increased variability in low mood (0.19, 95% CI: 0.07 to 0.31, p = 0.003) and in restless feelings (0.16, 95% CI: 0.03 to 0.28, p = 0.02) after 3 months of GAHT compared to baseline, and the increase sustains in variability in low mood after 12 months of GAHT compared to baseline (0.13; 95% CI: 0.01 to 0.26, p = 0.04). Absolute outcomes and analysis estimates are reported in Table 3.

### Group differences in changes in negative affect variability after starting GAHT

Compared to the TM group, we find a larger increase in variability in low mood (0.26, 95% CI: 0.09 to 0.42, p = 0.002), tense feelings (0.23, 95% CI: 0.03 to 0.43, p = 0.03) and restless feelings (0.33, 95% CI: 0.13 to 0.53, p = 0.002) in the TF group compared to TM group after 3 and 12 months of GAHT. All interaction analyses are reported in Table 3.

### Sensitivity analyses

Analyses excluding TF participants who started only anti-androgens or estrogens (n = 3), reported in Supplementary Table 3, shows similar results to the results reported in Table 3. Complete case analyses in the TM group, reported in Supplementary Tables 5 and 6, finds the same results as the overall results reported in Table 3. In the TF group, complete case analyses indicate an additional increase in variability of restless feelings after 12 months of feminizing GAHT. Addition of cycle regulation use (i.e., use of progestins or hormonal contraceptives) in the analyses in the TM group does not significantly change the direction or magnitude of the main results, as displayed in Supplementary Tables 7 and 8. At baseline, cycle regulation use is associated with lower variability in low mood, and in the longitudinal analyses cycle regulation use is also associated with lower variability in low mood. The use of psychotropic medication does not change the estimated group differences at baseline or changes over time in negative affect after starting GAHT, as displayed in Supplementary Tables 9 and 10, respectively.

## Discussion

The present study investigated changes in negative affect variability prospectively after 3 and 12 months of GAHT in transgender individuals, using four items in daily diaries. At baseline, TM individuals reported more variability in low mood compared to TF individuals. TM individuals reported a significant reduction in variability in low mood, tense feelings and restless feelings after 12 months of GAHT. In contrast, TF individuals reported an increase in variability in restless feelings after 3 months of GAHT, as well as an increase in variability in low mood after 3 months and 12 months of GAHT. Analyses comparing changes after starting GAHT between the TF and TM group indicated that the TF group shows significantly stronger increase in variability in low mood, tense feelings and restless feelings compared to the TM group.

Our results are in line with results from the study by Van Goozen et al. (1995), whose results show that after 3 months of GAHT TF participants reported higher daily ratings of “changeable mood” compared to TM participants. Furthermore, our results mirror the aforementioned sex differences in affect variability in cisgender populations: the TM participants report lower variability in low mood, tense feelings and restless feelings after 12 months of GAHT, in line with the lower affect variability in cisgender men, and the TF participants report higher variability in variability in restless feelings after 3 months of GAHT, as well as an increase in variability in low mood after 3 months and 12 months of GAHT, in line with higher variability in negative affect in cisgender women (Neiss & Almeida, 2004; Wang et al., 2012). Unfortunately, these previous studies did not report the means or standard deviations of their affect variability outcomes per sex, so we cannot directly compare our results to these previous results.

Our study findings in the TF group, who started using estrogens and anti-androgens, are in line with several findings in cisgender women and men. Sex hormone suppression through GnRH analogues or anti-androgens was found to increase affect variability in healthy cisgender women in reproductive age and cisgender men with prostate cancer, respectively (Donovan et al., 2015; Stenbæk et al., 2016). Sex hormone suppression combined with add-back of supraphysiological levels of estrogens and progestins was found to increase mood swings in cisgender women with a history of peripartum depression (Eisenlohr-Moul et al., 2024).

Results from covariate-adjusted analyses indicated that cycle regulation at baseline in TM participants was associated with lower variability in low mood, replicating the findings by Hamstra et al. (2017). We must interpret this result with caution, considering that some participants combined use of cycle regulation with testosterone, but nonetheless this replication shows that cycle regulation could also have an effect on affect variability.

In the interpretation of our findings, we must note that the size of the changes is relatively modest and we cannot ascertain that the found changes in mood variability would be noticeable in everyday life. Assessing effect sizes, we see that the largest group difference at baseline has a Cohen’s d effect size of 0.37 (i.e., difference in variability in sad mood), and changes in the TM and TF groups after starting GAHT result in Cohen’s d effect sizes of up to 0.51 (i.e., change in restless feelings after 12 months of masculinizing GAHT) and 0.45 (i.e., change in low mood after 3 months of feminizing GAHT). We would statistically consider these effect sizes as moderate, but future research should examine whether these changes also result in changes in affect dynamics noticeable in everyday life.

Although numerous explanations exist for changes in affect variability after starting GAHT, one potential explanation is based on neurobiological changes related to sex hormonal changes that occur in TM and TF individuals. Studies using emotion processing tasks (e.g. emotion reactivity) and neuroimaging (e.g. MRI and PET-imaging) suggest that sex hormones may impact neural processes involved in emotion processing, potentially through effects on serotonergic and GABA-ergic systems (Barth et al., 2015). Studies report increased serotonin transporter binding after 4 months of masculinizing GAHT and decreased binding following feminizing GAHT (Kranz et al., 2015).

Additionally, masculinizing GAHT has been found to increase monoamine oxidase-A levels after 4 to 9 months (Kranz et al., 2021), as well as to increase GABA-levels after an average of 5 months (Spurny-Dworak et al., 2022). Changes in these systems can influence emotional affect and processing (Frokjaer et al., 2008; Meyer et al., 2006), potentially impacting affect variability.

Additionally, psychosocial changes after starting GAHT likely contribute to our findings. Reductions in gender dysphoria and improvements in body image are commonly reported (van Leerdam et al., 2023), which can reduce emotional distress. However, previous findings show higher gender dysphoria scores after 6 months of GAHT in TF participants compared to TM participants, and improvements in social functioning after masculinizing but not feminizing GAHT (Foster Skewis et al., 2021), which implies that psychosocial effects of GAHT might also differ between the types of GAHT. Differences in emotional well-being could also be associated with TF persons experiencing more gender minority stress compared to TM persons (Poquiz et al., 2021).

Sociological research clearly shows that gender roles and stereotypes can influence one’s perception and reporting of emotions (Plant et al., 2000). Therefore, gendered expectations around emotion variability may also influence our results, as societal stereotypes often associate cisgender women with more “variable emotions” compared to cisgender men (Shields et al., 2018). Consequently, transgender persons may be more likely to endorse ‘’gender-stereotypical’’ emotional patterns. This tendency may further increase as physical changes align more closely with the desired gender after starting GAHT, although research on this topic is still scarce.

Within this study, one assumption we made is that participants were sufficiently able to identify and describe their emotions. An estimated 13 to 17% of people have difficulty identifying and describing their emotions, also known as alexithymia (Mason et al., 2005). Alexithymia is more prevalent in cisgender men compared to cisgender women (Levant et al., 2009), and a recent study indicates that alexithymia might be more prevalent in transgender persons compared to cisgender persons (Mazzoli et al., 2022). Interestingly, two recent studies found that alexithymia might reduce after starting GAHT, with Reed et al. (2023) finding reductions in alexithymia in TF persons after 4.5 months of GAHT and with Mazzoli et al. (2022) finding reductions in alexithymia in both TM and TF participants after 12 months of GAHT. Since being able to recognize and describe one’s emotions is essential to assess daily changes in affect, the presence of and possible changes in alexithymia after starting GAHT could have affected our results.

It should be noted that these biological and psychosocial explanations may intersect, rather than being mutually exclusive. Changes in social environments after GAHT initiation, resulting in reductions in mental and social distress (Collet et al., 2023), could affect emotion-regulating systems in the brain. Vice versa, neurobiological changes after starting GAHT could affect emotion processing and recognition of emotions (Reed et al., 2023), which could influence coping with emotions in relation to the social environment. Thus, our results serve as descriptive insights into reported changes in negative affect variability after GAHT use, but further research is needed to disentangle underlying mechanisms and causality.

This is one of the first prospective studies on type of GAHT (e.g. masculinizing and feminizing) and variability in day-to-day negative affect during the first year of GAHT use. We deem the approach we took in this study, examining direct day-to-day variations in negative effect using diary data, as one of the strengths of this study. The methodologies used in previous studies on sex hormones and affect exhibit considerable variation. Some studies use retrospective questionnaires inquiring about depression or affect (Aldridge et al., 2021; Morssinkhof et al., 2024), a method susceptible to recall bias, while others rely on the mean of daily self-reports (Donovan et al., 2015; Hamstra et al., 2017; Van Goozen et al., 1995), which introduces potential bias by mistakenly reflecting average effects rather than true variability. Since we used daily diaries during one week, our measurements are likely to be less subject to recall bias and could most presumably better account for day-to-day variations in negative affect, although this approach might still not be optimal. Additionally, we corrected negative affect variability changes for average negative affect, which is a methodological improvement to better account for the influence of possible floor effects in our participants’ mean affect (Maciejewski et al., 2023).

Although this study enabled us to prospectively examine the influences of sex hormones on negative affect variability, there are also several limitations in our setup. First, we used four self-formulated outcomes to measure negative affect. Although these symptoms are based on key symptoms for depressive and anxiety disorders, they are not validated for the purpose of studying negative affect variability. This emphasizes the need for future studies to examine affect variability using validated affect scales, for instance based on the PANAS (Positive and Negative Affect Schedule; (Watson et al., 1988) or POMS (Profile of Mood States; (McNair D, 1992).

Second, we did not examine positive affect items alongside negative affect items. Previous literature in cisgender samples shows that those who report more intense negative affect also tend to report more intense positive affect, and vice versa (Fujita et al., 1991; Larsen & Diener, 1987). Similarly, individuals who report stronger variability in positive affect also tend to report stronger variability in negative affect (Larsen & Diener, 1987). Matthys et al. (2021) found changes in both positive and negative affect in the first 18 months of GAHT, but the authors did not assess affect variability. However, this emphasizes the relevance of also assessing positive affect changes after starting GAHT.

Third, our study has measured variations in negative affect from day to day but could not capture fluctuations within a single day. To this point, one could also debate whether the variability reported in this paper should be called “affect variability” or “mood variability”, with mood variability suggesting a longer timeframe. Retrospective reporting of emotions over a whole day has been shown to be less reliable than reported momentary emotions over the last hours (Solhan et al., 2009), and therefore the longer timeframe used in this study is a limitation.

Last, contextual information on daily events which may influence negative affect, including presence or absence of social support, gender affirmation by others or other contextual factors, are lacking in our study. This is especially important since our study examines changes after starting GAHT. Starting GAHT is associated with a plethora of psychological and social changes, including reduced gender dysphoria, improved body image, as well as improvements in psychosocial functioning (Doyle et al., 2023; Foster Skewis et al., 2021; van Leerdam et al., 2023).

Future studies on affect variability in transgender people or in the context of sex hormones should focus on using validated affect items to study affect variability, including positive affect. This should preferably be done using non-invasive technologies such as Ecological Momentary Assessment (EMA) (Shiffman et al., 2008), which enables researchers to inquire about affect for up to eight times per day. Using tools such as EMA measurements can further enable researchers to account for contextual factors, which can provide further insights into the possible causes and consequences of changes in affect variability.

In summary, our findings suggest that variability in affect in transgender persons can shift from a pattern consistent with sex assigned at birth before GAHT towards a pattern more in line with gender identity after GAHT. However, the results should be interpreted carefully, since the demonstrated changes in affect variability are modest and contextual information on daily events is lacking in our study. Since affect variability is a key feature of one’s emotional experiences, future studies should provide more information on affect variability changes after GAHT and on possible underlying mechanisms, with measurement of both negative and positive affect variability within social and daily contexts during GAHT.

## CRediT statement

**Margot W. L. Morssinkhof:** Conceptualization, Methodology, Formal analysis, Investigation, Data Curation, Writing – Original Draft. **Marijn Schipper:** Conceptualization, Methodology, Formal analysis, Writing – Original Draft. **Baudewijntje P. C. Kreukels:** Conceptualization, Writing – Review & Editing; **Karin van der Tuuk:** Resources, Investigation; **Martin den Heijer:** Resources, Investigation, Writing – Review & Editing; **Odile A. van den Heuvel:** Conceptualization, Investigation, Writing – Review & Editing; **David Matthew Doyle:** Conceptualization, Formal Analysis, Writing – Review & Editing, Supervision, Funding Acquisition; **Birit F. P. Broekman:** Conceptualization, Investigation, Writing – Review & Editing, Supervision, Funding Acquisition.

## Data Availability

Since data of this study contains patient data from a vulnerable group (i.e., transgender participants), anonymized study data is available upon reasonable request by contacting the corresponding author. The study codebook and analysis code for all analyses is available via osf.io/8dnjp/.

## Acknowledgements

We are grateful to the research participants of the RESTED study, whose participation was essential for this research.

## Supplementary materials

### 1. iMean and Mean Absolute Successive Difference

#### iMean

Supplementary Table 1 displays the iMean per participant group and timepoint. At baseline, the TF group shows significantly lower mean levels of restless feelings compared to the TM group (-0.33, 95% CI: -0.64 to -0.02, p = 0.04), but there are no significant group differences in means of the other outcomes. Stratified linear mixed models in the TM group indicate that means of low mood (0.13, 95% CI: 0.00 to 0.25, p = 0.04), less interest (0.14, 95% CI: 0.04 to 0.26, p = 0.01) and restless feelings (0.21, 95% CI: 0.07 to 0.35, p = 0.003) increase after 12 months of GAHT, while the means of tense feelings do not significantly change after GAHT. Stratified analyses in the TF group show no significant changes in means of low mood, less interest, tense feelings or restless feelings. Compared to the TM group, the TF group shows a smaller increase in mean less interest (-0.19, 95% CI: -0.36 to - 0.02, p = 0.03) and restless feelings (-0.21, 95% CI: 0.40 to -0.01, p = 0.04) after 12 months of GAHT as reported in Supplementary Table 1.

#### MASD

We reported the Mean Absolute Successive Difference (MASD) in Supplementary Tables 1 and 2. At baseline, the MASD outcomes show lower variability for low mood in the TF group compared to the TM group (-0.16; 95% CI: -0.28 to -0.04, p = 0.01). Stratified analyses on the MASD find a decrease in variability of restless feelings in the TM group after 12 months of GAHT (-0.26, 95% CI: -0.43 to -0.09, p = 0.004), whereas the MASD of low mood increases in the TF group after 3 and 12 months of GAHT (0.16, 95% CI: 0.03 to 0.28, p = 0.02; 0.14, 95% CI: 0.01 to 0.27, p = 0.04). We find a significant group difference in low mood (0.24, 95% CI: 0.06 to 0.42, p = 0.01) and restless feelings (0.35, 95% CI: 0.13 to 0.57, p = 0.002), with the TF group reporting larger increases in variability for both items after 3 and 12 months of GAHT compared to the TM group. Altogether, results show similar directions to the iSD outcomes, but for some items, the iSD changes significantly whereas the MASD does not.

**Supplementary Table 1.**
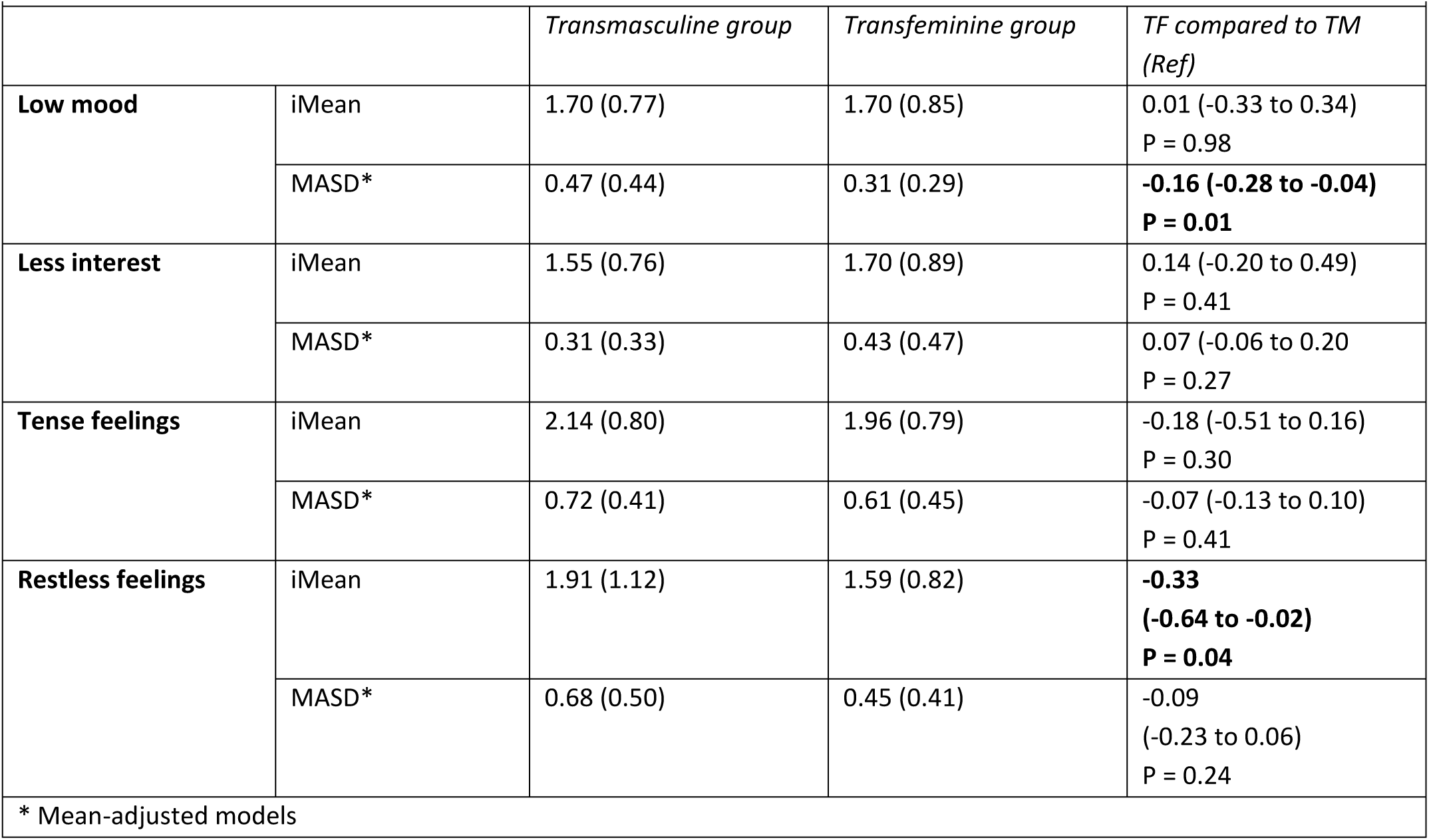
iMeans and mean absolute successive difference (MASD) scores in the negative affect outcomes at baseline.

**Supplementary Table 2.**
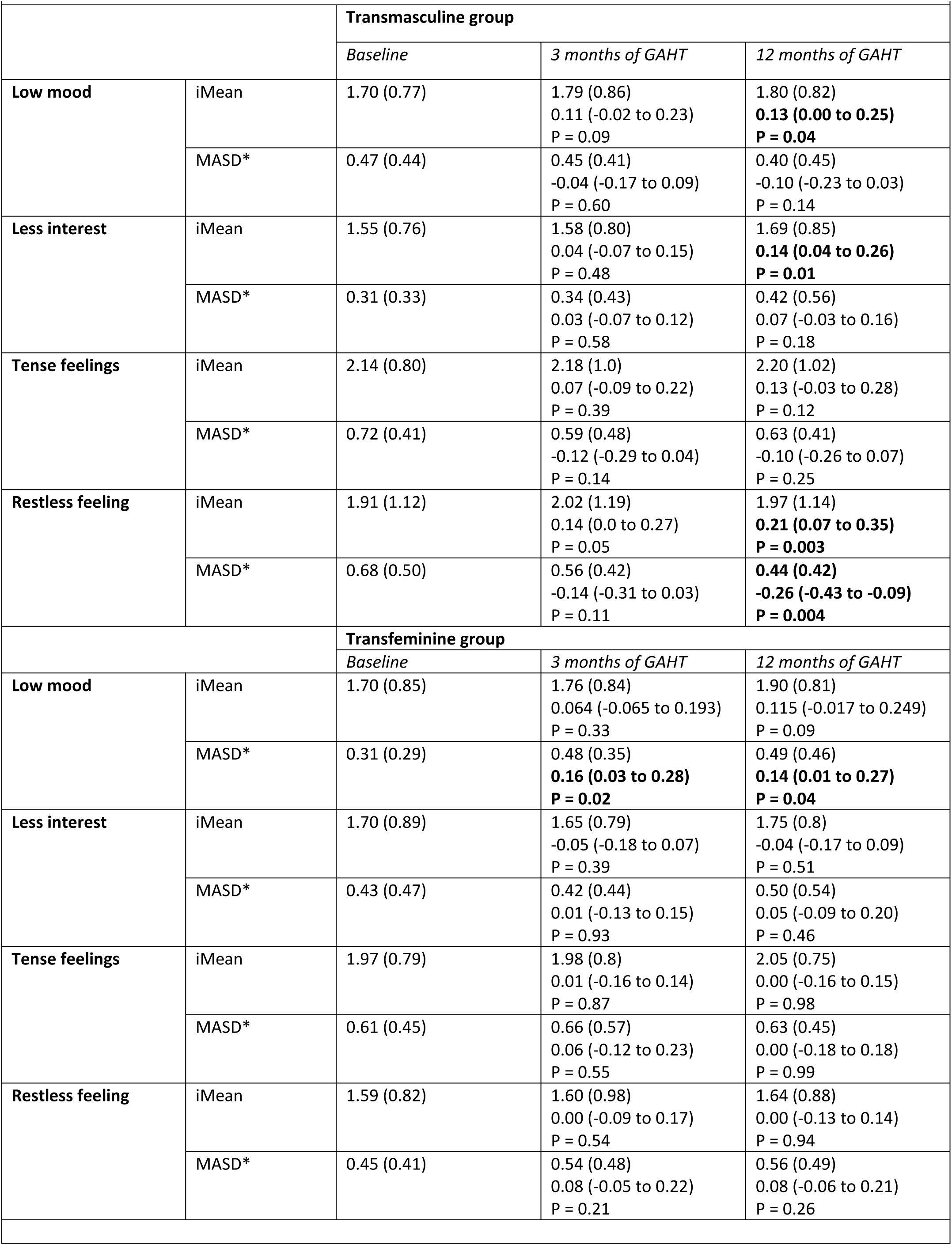
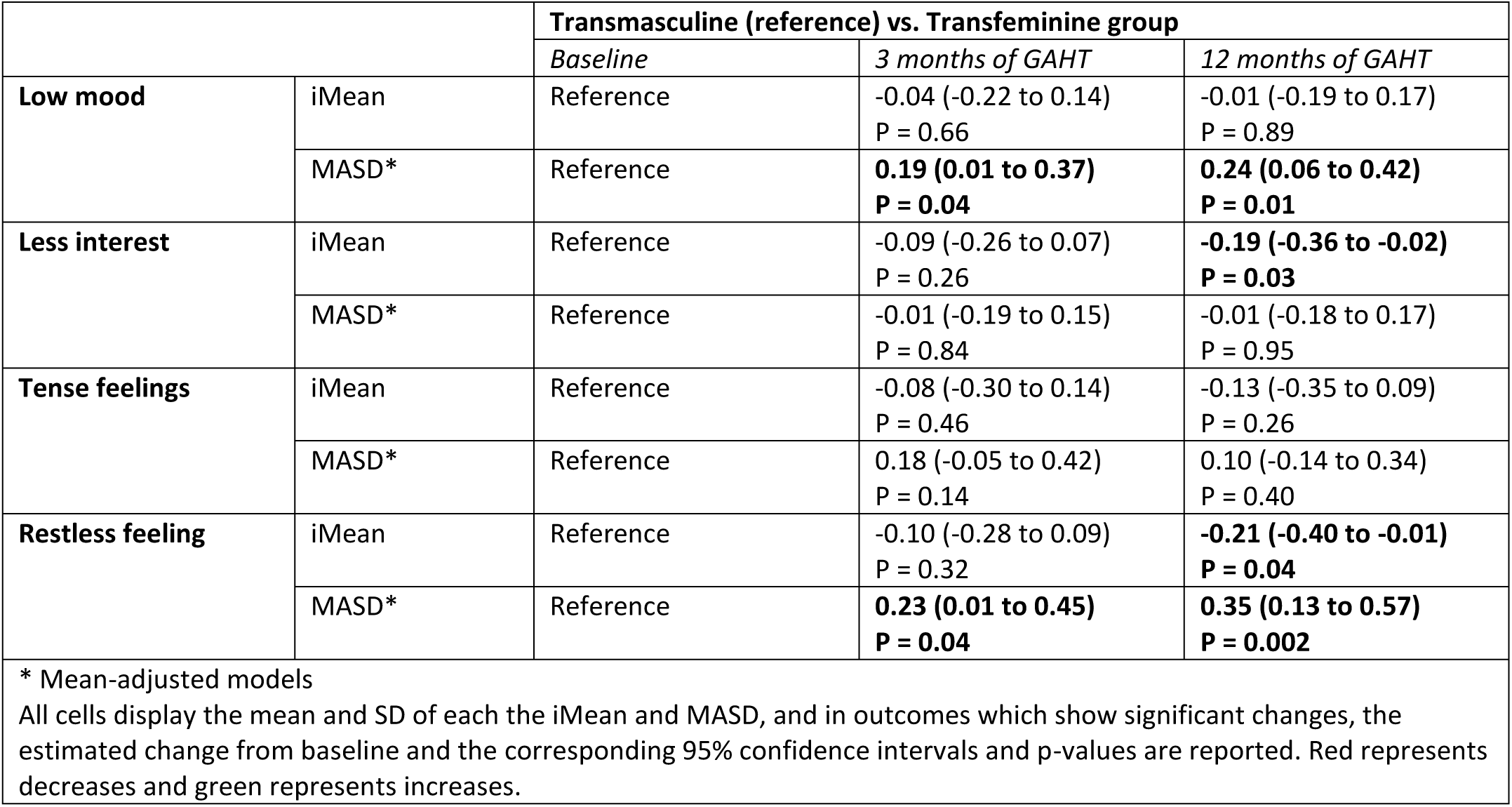
Changes in iMeans and MASD after GAHT use, analyses stratified per group.

### 2. Only combined feminizing GAHT (i.e., anti-androgens and estrogens)

Supplementary Table 3 displays the results of analyses excluding TF participants who only used anti-androgens or estrogens; the resulting sample consists of 42 TF participants.

**Supplementary Table 3.**
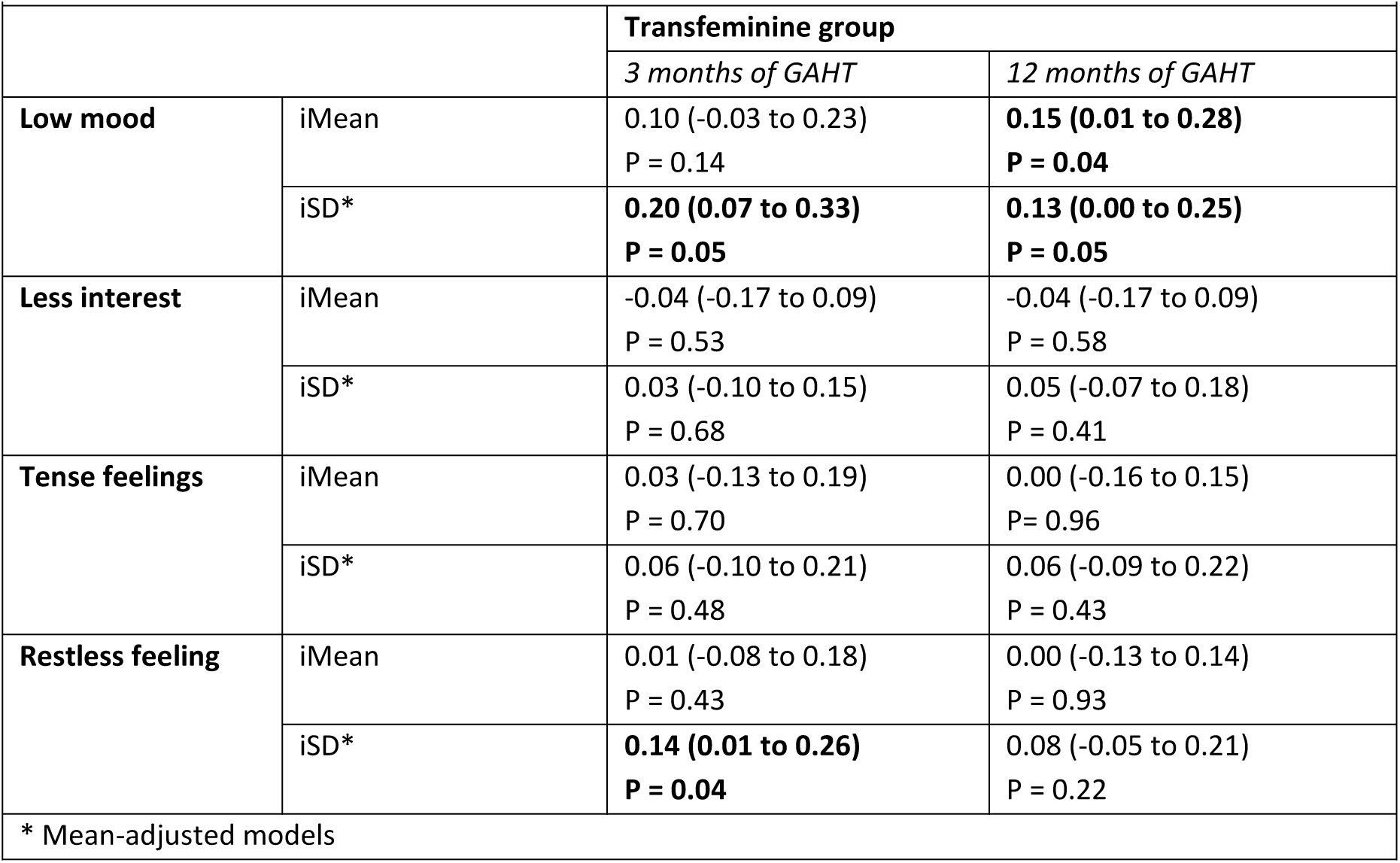
Changes in negative affect after GAHT use, stratified per group. All cells display the mean and SD of each iMean and iSD, and in outcomes which show significant changes, the estimated change from baseline and the corresponding 95% confidence intervals and p-values are reported. Red represents decreases and green represents increases.

### 3. Loss to follow-up & complete case analysis

In Supplementary Table 4, we display the baseline demographic characteristics of participants who were and were not lost to follow-up.

**Supplementary Table 4.**
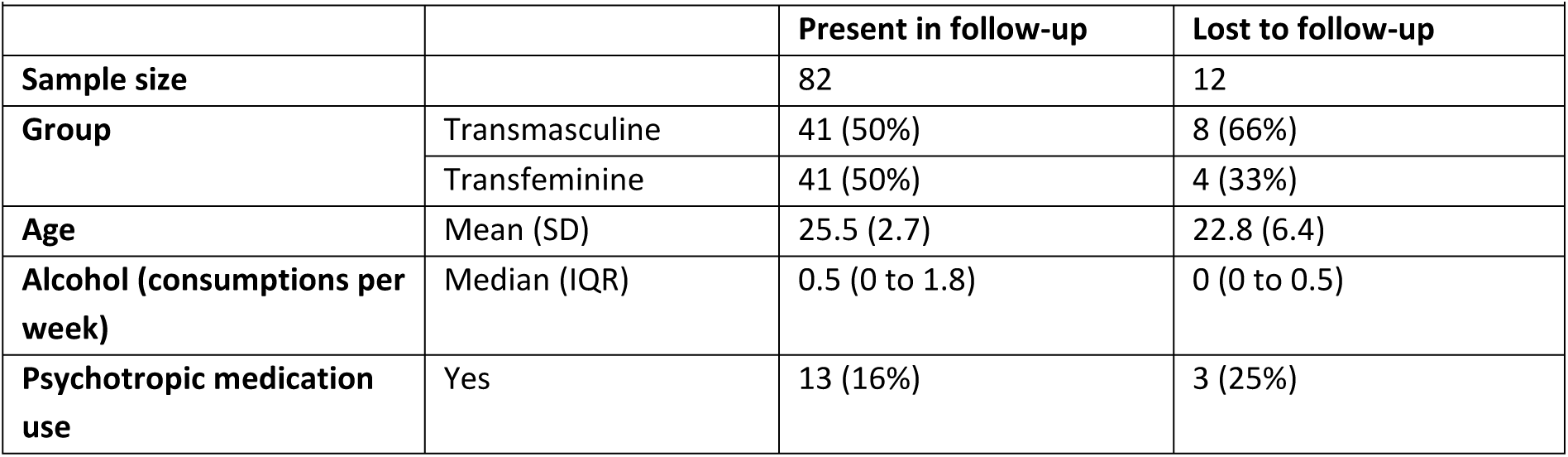
Demographic characteristics for participants present in follow-up compared to lost to follow-up.

Supplementary Tables 5 and 6 display complete case analyses, i.e. analyses including only participants who contributed both a baseline and at least one follow-up measurement. Overall results show similar directions and magnitudes compared to the main results.

**Supplementary Table 5.**
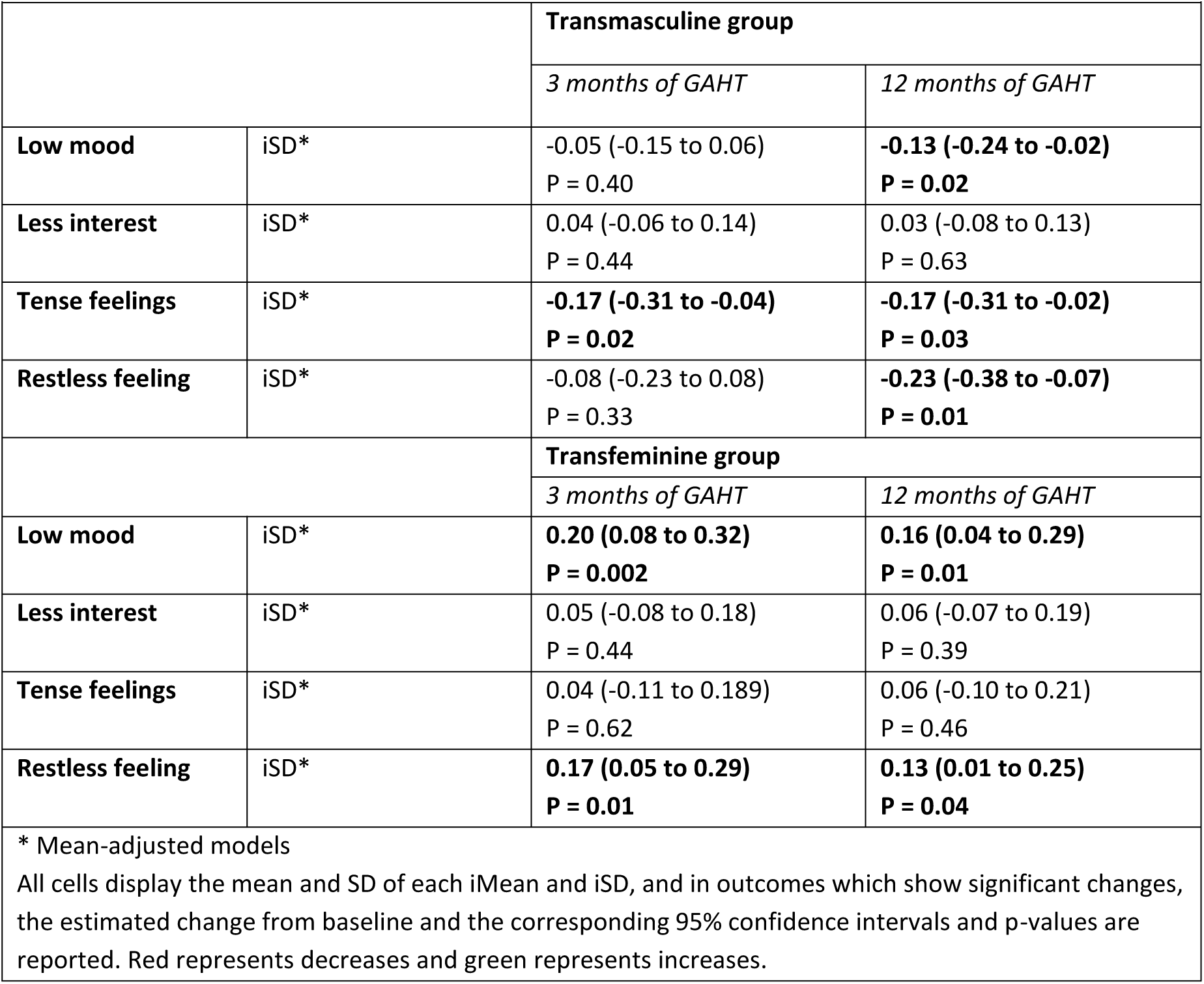
Changes in negative affect after GAHT use, stratified per group.

**Supplementary Table 6.**
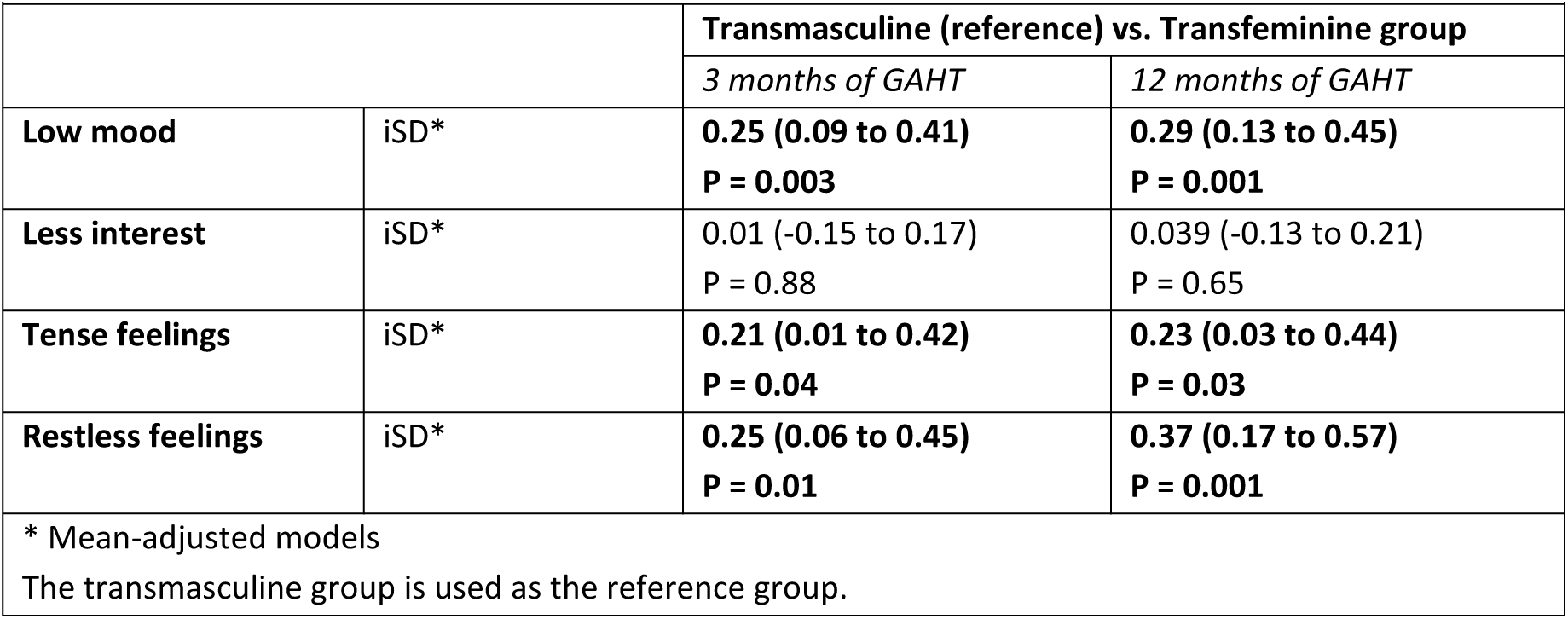
Changes in negative affect outcomes after GAHT use, interaction analyses.

### 4. Cycle regulation

In Supplementary Table 7, we display differences between the TF group and the TM group on and off cycle regulation at baseline. The results show that the both groups (i.e., TM group with and without cycle regulation) have higher variability in low mood and restless feelings compared to the TF group at baseline.

**Supplementary Table 7.**
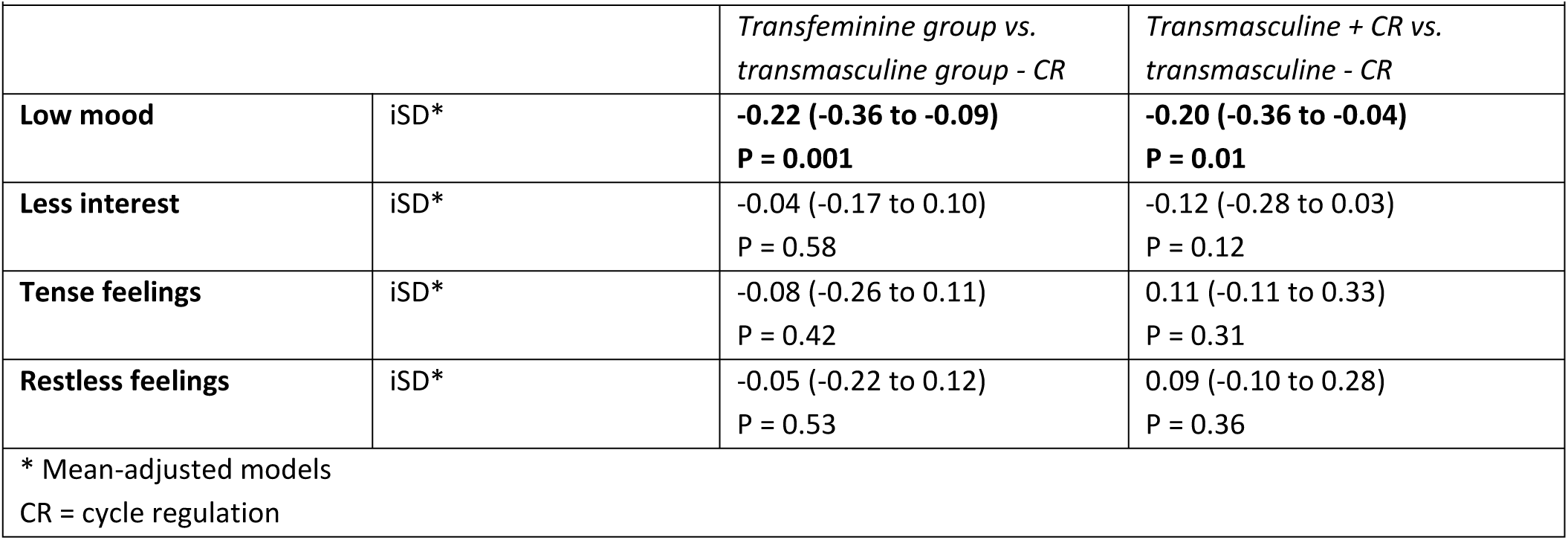
Negative affect outcomes before GAHT use (**baseline**), examining group differences and adjusted for cycle regulation use and psychotropic medication use. The transmasculine group not using cycle regulation is used as the reference group.

In Supplementary Table 8, we display the changes in affect variability after correction for cycle regulation in the TM group. The results overall show similar directions and magnitude compared to the results without correction for cycle regulation, although cycle regulation is overall associated with lower variability in low mood.

**Supplementary Table 8.**
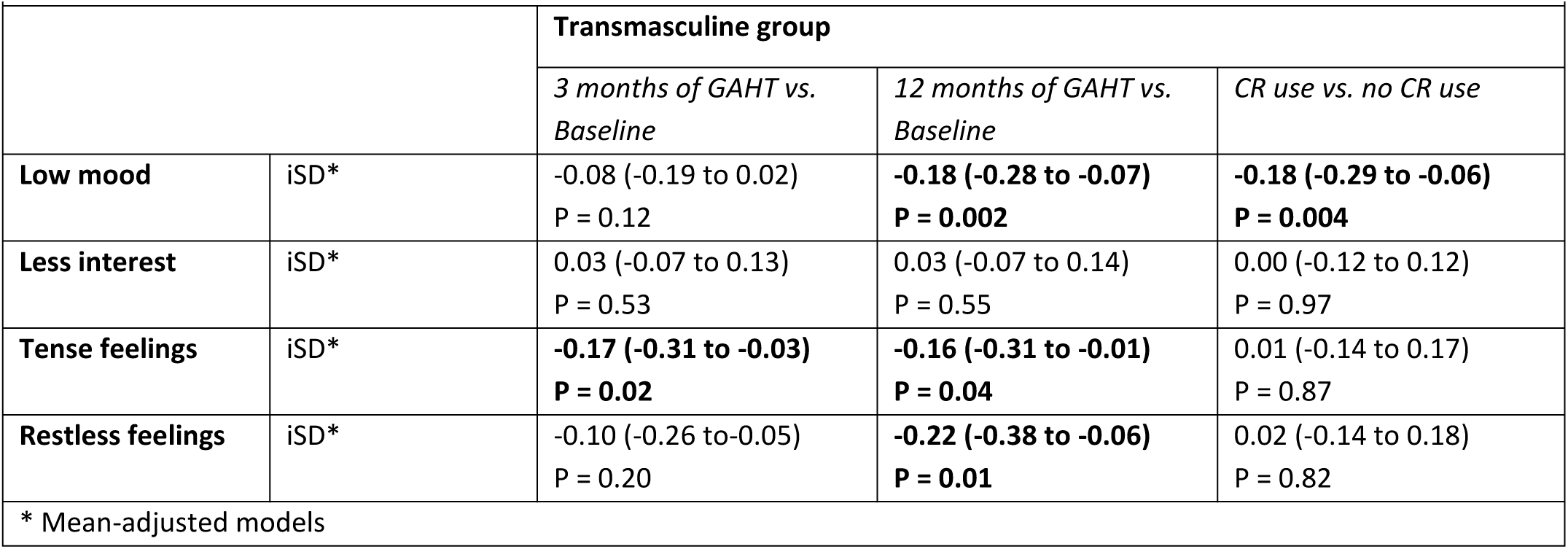
Changes in negative affect after GAHT use, stratified per group and adjusted for cycle regulation and psychotropic medication use. The estimated change from baseline and the corresponding 95% confidence intervals and p-values are reported.

### 5. Psychotropic medication

Psychotropic medication was added as covariate to the model – main effects and adjusted group effects at baseline. The longitudinal analyses in the TM group indicate that psychotropic medication use is associated with more variable feelings of less interest. In the longitudinal analyses in the TF group, psychotropic medication use is associated with less variable low mood, less variable tense feelings, and less variable restless feelings. Results are displayed in Supplementary Table 9.

**Supplementary Table 9.**
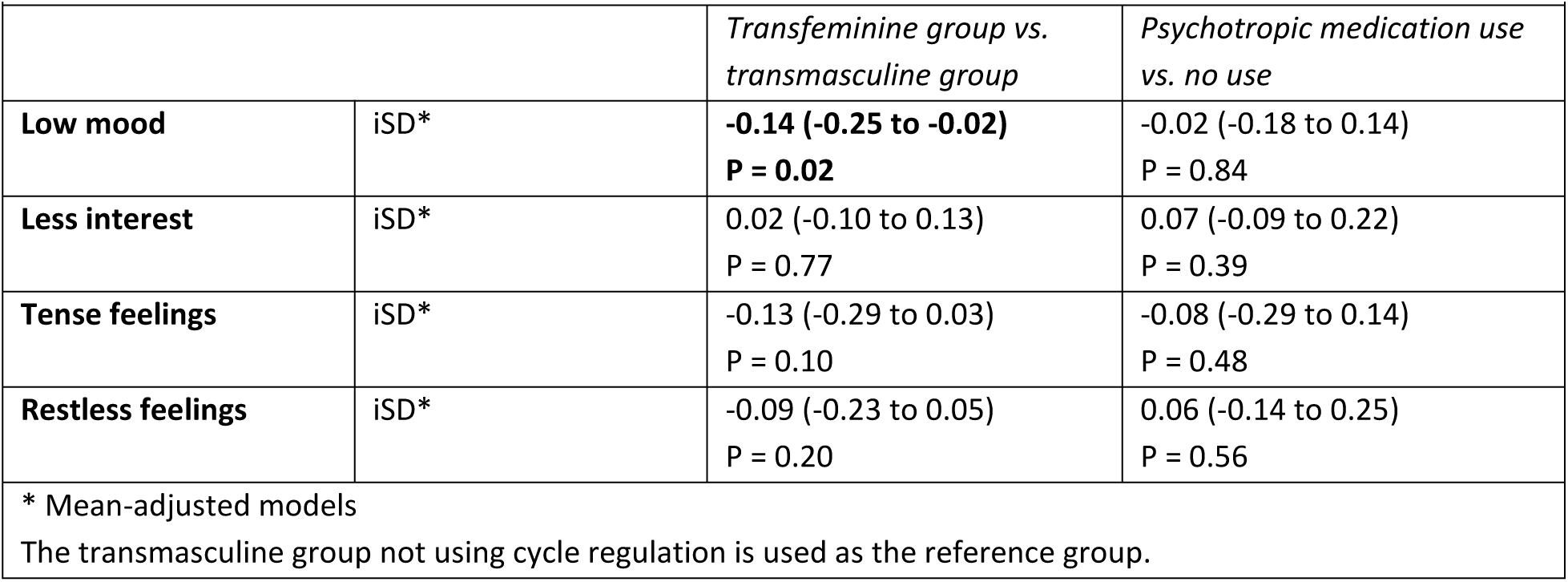
Negative affect outcomes before GAHT use (baseline), examining group differences and adjusted for cycle regulation use and psychotropic medication use.

Supplementary Table 10 shows the analyses with the addition of psychotropic medication as a covariate: addition of this covariate does not change the main results.

**Supplementary Table 10.**
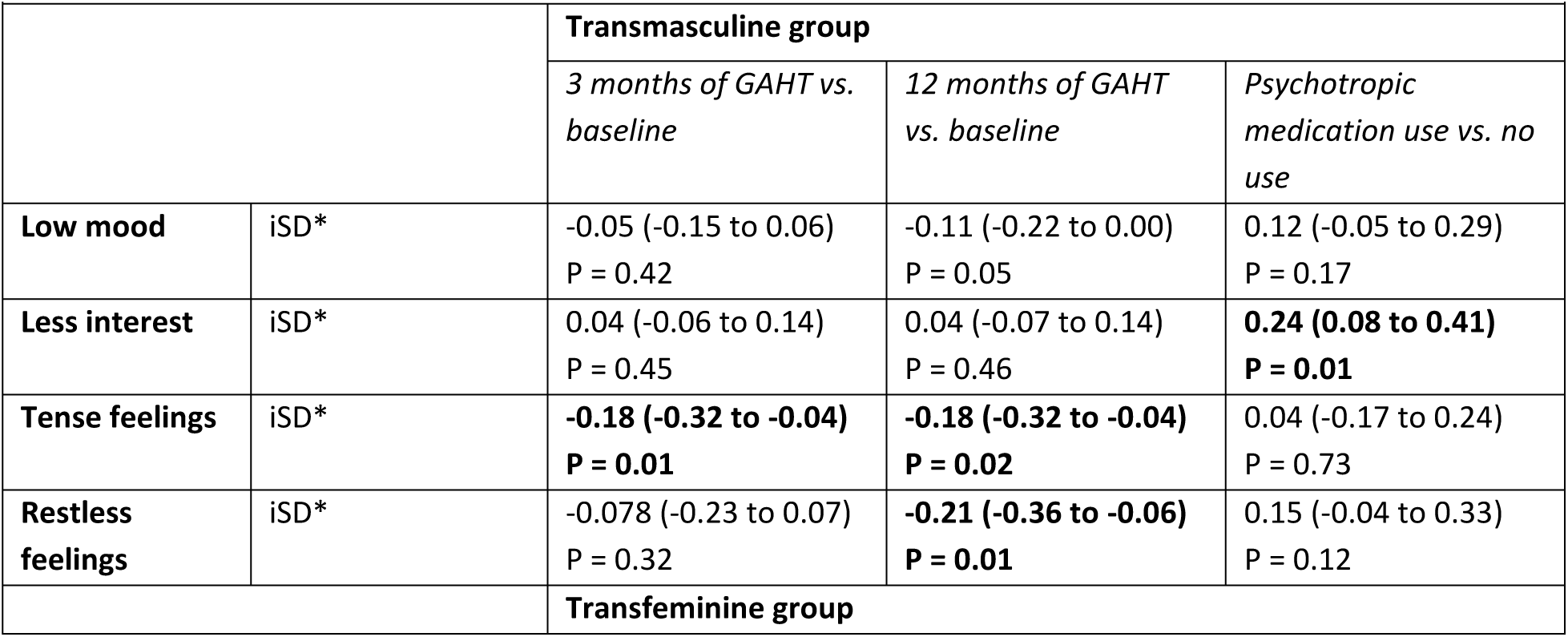
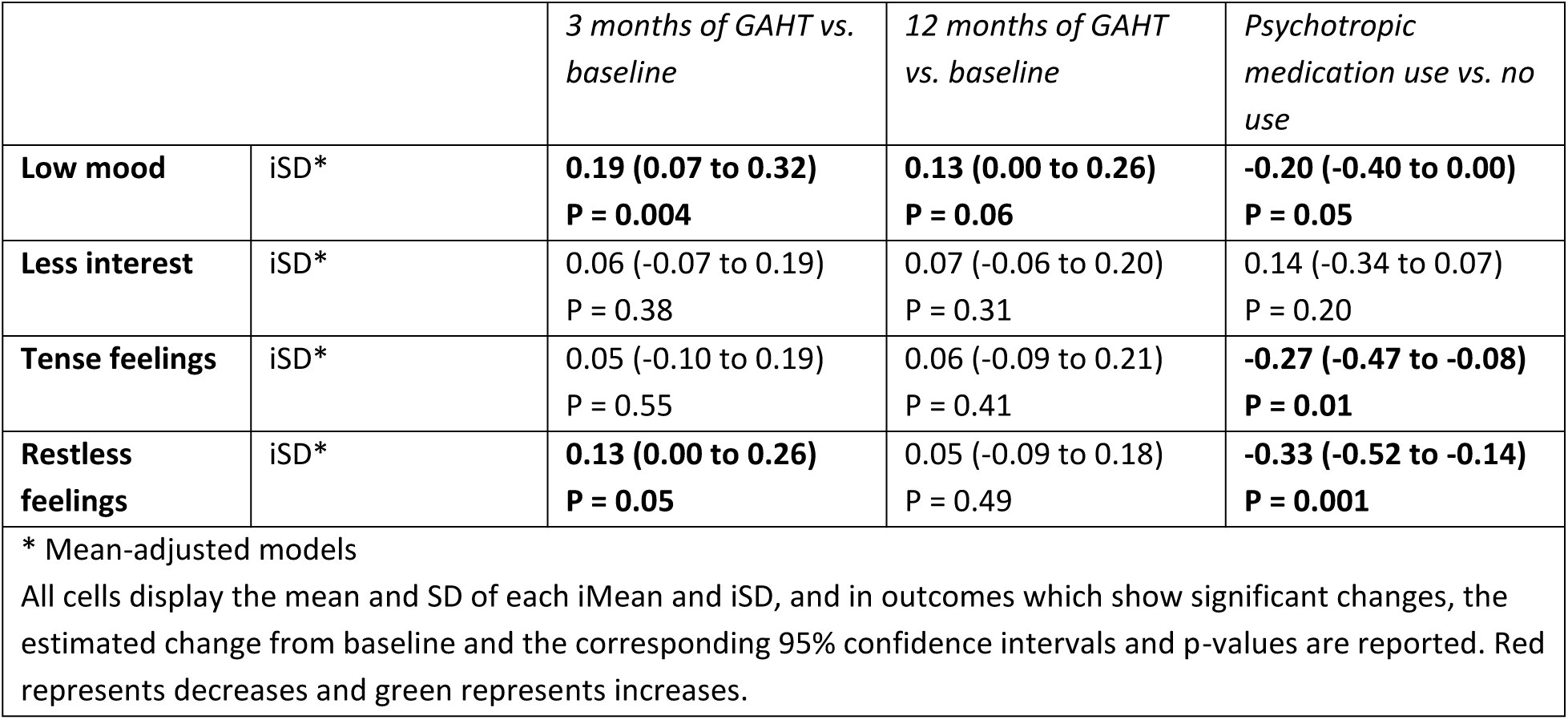
Changes in negative affect after GAHT use, stratified per group and adjusted for cycle regulation and psychotropic medication use.

