## Supplementary Materials for "Changes in affect variability after starting gender-affirming hormone therapy"

| **Supplementary Table 1**. iMeans and mean absolute successive difference (MASD) scores in the negative affect outcomes at baseline. | | | | |
| --- | --- | --- | --- | --- |
|  | | *Transmasculine group* | *Transfeminine group* | *TF compared to TM (Ref)* |
| **Low mood** | iMean | 1.70 (0.77) | 1.70 (0.85) | 0.01 (-0.33 to 0.34)  P = 0.98 |
|  | MASD* | 0.47 (0.44) | 0.31 (0.29) | **-0.16 (-0.28 to -0.04)**  **P = 0.01** |
| **Less interest** | iMean | 1.55 (0.76) | 1.70 (0.89) | 0.14 (-0.20 to 0.49)  P = 0.41 |
|  | MASD* | 0.31 (0.33) | 0.43 (0.47) | 0.07 (-0.06 to 0.20  P = 0.27 |
| **Tense feelings** | iMean | 2.14 (0.80) | 1.96 (0.79) | -0.18 (-0.51 to 0.16)  P = 0.30 |
|  | MASD* | 0.72 (0.41) | 0.61 (0.45) | -0.07 (-0.13 to 0.10)  P = 0.41 |
| **Restless feelings** | iMean | 1.91 (1.12) | 1.59 (0.82) | **-0.33**  **(-0.64 to -0.02)**  **P = 0.04** |
|  | MASD* | 0.68 (0.50) | 0.45 (0.41) | -0.09  (-0.23 to 0.06)  P = 0.24 |
| * Mean-adjusted models | | | | |

| **Supplementary Table 2.** Changes in iMeans and MASD after GAHT use, analyses stratified per group. | | | | |
| --- | --- | --- | --- | --- |
|  | | **Transmasculine group** | | |
|  |  | *Baseline* | *3 months of GAHT* | *12 months of GAHT* |
| **Low mood** | iMean | 1.70 (0.77) | 1.79 (0.86)  0.11 (-0.02 to 0.23)  P = 0.09 | 1.80 (0.82)  **0.13 (0.00 to 0.25) P = 0.04** |
|  | MASD* | 0.47 (0.44) | 0.45 (0.41)  -0.04 (-0.17 to 0.09)  P = 0.60 | 0.40 (0.45)  -0.10 (-0.23 to 0.03)  P = 0.14 |
| **Less interest** | iMean | 1.55 (0.76) | 1.58 (0.80)  0.04 (-0.07 to 0.15)  P = 0.48 | 1.69 (0.85)  **0.14 (0.04 to 0.26)**  **P = 0.01** |
|  | MASD* | 0.31 (0.33) | 0.34 (0.43)  0.03 (-0.07 to 0.12)  P = 0.58 | 0.42 (0.56)  0.07 (-0.03 to 0.16)  P = 0.18 |
| **Tense feelings** | iMean | 2.14 (0.80) | 2.18 (1.0)  0.07 (-0.09 to 0.22)  P = 0.39 | 2.20 (1.02)  0.13 (-0.03 to 0.28)  P = 0.12 |
|  | MASD* | 0.72 (0.41) | 0.59 (0.48)  -0.12 (-0.29 to 0.04)  P = 0.14 | 0.63 (0.41)  -0.10 (-0.26 to 0.07)  P = 0.25 |
| **Restless feeling** | iMean | 1.91 (1.12) | 2.02 (1.19)  0.14 (0.0 to 0.27)  P = 0.05 | 1.97 (1.14)  **0.21 (0.07 to 0.35)**  **P = 0.003** |
|  | MASD* | 0.68 (0.50) | 0.56 (0.42)  -0.14 (-0.31 to 0.03)  P = 0.11 | **0.44 (0.42)**  **-0.26 (-0.43 to -0.09)**  **P = 0.004** |
|  | | **Transfeminine group** | | |
|  |  | *Baseline* | *3 months of GAHT* | *12 months of GAHT* |
| **Low mood** | iMean | 1.70 (0.85) | 1.76 (0.84)  0.064 (-0.065 to 0.193)  P = 0.33 | 1.90 (0.81)  0.115 (-0.017 to 0.249)  P = 0.09 |
|  | MASD* | 0.31 (0.29) | 0.48 (0.35)  **0.16 (0.03 to 0.28)**  **P = 0.02** | 0.49 (0.46)  **0.14 (0.01 to 0.27)**  **P = 0.04** |
| **Less interest** | iMean | 1.70 (0.89) | 1.65 (0.79)  -0.05 (-0.18 to 0.07)  P = 0.39 | 1.75 (0.8)  -0.04 (-0.17 to 0.09)  P = 0.51 |
|  | MASD* | 0.43 (0.47) | 0.42 (0.44)  0.01 (-0.13 to 0.15)  P = 0.93 | 0.50 (0.54)  0.05 (-0.09 to 0.20)  P = 0.46 |
| **Tense feelings** | iMean | 1.97 (0.79) | 1.98 (0.8)  0.01 (-0.16 to 0.14)  P = 0.87 | 2.05 (0.75)  0.00 (-0.16 to 0.15)  P = 0.98 |
|  | MASD* | 0.61 (0.45) | 0.66 (0.57)  0.06 (-0.12 to 0.23)  P = 0.55 | 0.63 (0.45)  0.00 (-0.18 to 0.18)  P = 0.99 |
| **Restless feeling** | iMean | 1.59 (0.82) | 1.60 (0.98)  0.00 (-0.09 to 0.17)  P = 0.54 | 1.64 (0.88)  0.00 (-0.13 to 0.14)  P = 0.94 |
|  | MASD* | 0.45 (0.41) | 0.54 (0.48)  0.08 (-0.05 to 0.22)  P = 0.21 | 0.56 (0.49)  0.08 (-0.06 to 0.21)  P = 0.26 |
|  | | **Transmasculine (reference) vs. Transfeminine group** | | |
|  |  | *Baseline* | *3 months of GAHT* | *12 months of GAHT* |
| **Low mood** | iMean | Reference | -0.04 (-0.22 to 0.14)  P = 0.66 | -0.01 (-0.19 to 0.17)  P = 0.89 |
|  | MASD* | Reference | **0.19 (0.01 to 0.37)**  **P = 0.04** | **0.24 (0.06 to 0.42)**  **P = 0.01** |
| **Less interest** | iMean | Reference | -0.09 (-0.26 to 0.07)  P = 0.26 | **-0.19 (-0.36 to -0.02)**  **P = 0.03** |
|  | MASD* | Reference | -0.01 (-0.19 to 0.15)  P = 0.84 | -0.01 (-0.18 to 0.17)  P = 0.95 |
| **Tense feelings** | iMean | Reference | -0.08 (-0.30 to 0.14)  P = 0.46 | -0.13 (-0.35 to 0.09)  P = 0.26 |
|  | MASD* | Reference | 0.18 (-0.05 to 0.42)  P = 0.14 | 0.10 (-0.14 to 0.34)  P = 0.40 |
| **Restless feeling** | iMean | Reference | -0.10 (-0.28 to 0.09)  P = 0.32 | **-0.21 (-0.40 to -0.01)**  **P = 0.04** |
|  | MASD* | Reference | **0.23 (0.01 to 0.45)**  **P = 0.04** | **0.35 (0.13 to 0.57)**  **P = 0.002** |
| * Mean-adjusted models  All cells display the mean and SD of each the iMean and MASD, and in outcomes which show significant changes, the estimated change from baseline and the corresponding 95% confidence intervals and p-values are reported. Red represents decreases and green represents increases. | | | | |

### 2. Only combined feminizing GAHT (i.e., anti-androgens and estrogens)

Supplementary Table 3 displays the results of analyses excluding TF participants who only used anti-androgens or estrogens; the resulting sample consists of 42 TF participants.

| **Supplementary Table 3**. Changes in negative affect after GAHT use, stratified per group. All cells display the mean and SD of each iMean and iSD, and in outcomes which show significant changes, the estimated change from baseline and the corresponding 95% confidence intervals and p-values are reported. Red represents decreases and green represents increases. | | | |
| --- | --- | --- | --- |
|  | | **Transfeminine group** | |
|  |  | *3 months of GAHT* | *12 months of GAHT* |
| **Low mood** | iMean | 0.10 (-0.03 to 0.23)  P = 0.14 | **0.15 (0.01 to 0.28)**  **P = 0.04** |
|  | iSD* | **0.20 (0.07 to 0.33)**  **P = 0.05** | **0.13 (0.00 to 0.25)**  **P = 0.05** |
| **Less interest** | iMean | -0.04 (-0.17 to 0.09)  P = 0.53 | -0.04 (-0.17 to 0.09)  P = 0.58 |
|  | iSD* | 0.03 (-0.10 to 0.15)  P = 0.68 | 0.05 (-0.07 to 0.18)  P = 0.41 |
| **Tense feelings** | iMean | 0.03 (-0.13 to 0.19)  P = 0.70 | 0.00 (-0.16 to 0.15)  P= 0.96 |
|  | iSD* | 0.06 (-0.10 to 0.21)  P = 0.48 | 0.06 (-0.09 to 0.22)  P = 0.43 |
| **Restless feeling** | iMean | 0.01 (-0.08 to 0.18)  P = 0.43 | 0.00 (-0.13 to 0.14)  P = 0.93 |
|  | iSD* | **0.14 (0.01 to 0.26)**  **P = 0.04** | 0.08 (-0.05 to 0.21)  P = 0.22 |
| * Mean-adjusted models | | | |

### 3. Loss to follow-up & complete case analysis

In Supplementary Table 4, we display the baseline demographic characteristics of participants who were and were not lost to follow-up.

| **Supplementary Table 4.** Demographic characteristics for participants present in follow-up compared to lost to follow-up. | | | |
| --- | --- | --- | --- |
|  |  | **Present in follow-up** | **Lost to follow-up** |
| **Sample size** |  | 82 | 12 |
| **Group** | Transmasculine | 41 (50%) | 8 (66%) |
|  | Transfeminine | 41 (50%) | 4 (33%) |
| **Age** | Mean (SD) | 25.5 (2.7) | 22.8 (6.4) |
| **Alcohol (consumptions per week)** | Median (IQR) | 0.5 (0 to 1.8) | 0 (0 to 0.5) |
| **Psychotropic medication use** | Yes | 13 (16%) | 3 (25%) |

Supplementary Tables 5 and 6 display complete case analyses, i.e. analyses including only participants who contributed both a baseline and at least one follow-up measurement. Overall results show similar directions and magnitudes compared to the main results.

| **Supplementary Table 5.** Changes in negative affect after GAHT use, stratified per group. | | | |
| --- | --- | --- | --- |
|  | | **Transmasculine group** | |
|  |  | *3 months of GAHT* | *12 months of GAHT* |
| **Low mood** | iSD* | -0.05 (-0.15 to 0.06)  P = 0.40 | **-0.13 (-0.24 to -0.02)**  **P = 0.02** |
| **Less interest** | iSD* | 0.04 (-0.06 to 0.14)  P = 0.44 | 0.03 (-0.08 to 0.13)  P = 0.63 |
| **Tense feelings** | iSD* | **-0.17 (-0.31 to -0.04)**  **P = 0.02** | **-0.17 (-0.31 to -0.02)**  **P = 0.03** |
| **Restless feeling** | iSD* | -0.08 (-0.23 to 0.08)  P = 0.33 | **-0.23 (-0.38 to -0.07)**  **P = 0.01** |
|  | | **Transfeminine group** | |
|  |  | *3 months of GAHT* | *12 months of GAHT* |
| **Low mood** | iSD* | **0.20 (0.08 to 0.32)**  **P = 0.002** | **0.16 (0.04 to 0.29)**  **P = 0.01** |
| **Less interest** | iSD* | 0.05 (-0.08 to 0.18)  P = 0.44 | 0.06 (-0.07 to 0.19)  P = 0.39 |
| **Tense feelings** | iSD* | 0.04 (-0.11 to 0.189)  P = 0.62 | 0.06 (-0.10 to 0.21)  P = 0.46 |
| **Restless feeling** | iSD* | **0.17 (0.05 to 0.29)**  **P = 0.01** | **0.13 (0.01 to 0.25)**  **P = 0.04** |
| * Mean-adjusted models  All cells display the mean and SD of each iMean and iSD, and in outcomes which show significant changes, the estimated change from baseline and the corresponding 95% confidence intervals and p-values are reported. Red represents decreases and green represents increases. | | | |

| **Supplementary Table 6.** Changes in negative affect outcomes after GAHT use, interaction analyses. | | | |
| --- | --- | --- | --- |
|  | | **Transmasculine (reference) vs. Transfeminine group** | |
|  |  | *3 months of GAHT* | *12 months of GAHT* |
| **Low mood** | iSD* | **0.25 (0.09 to 0.41)**  **P = 0.003** | **0.29 (0.13 to 0.45)**  **P = 0.001** |
| **Less interest** | iSD* | 0.01 (-0.15 to 0.17)  P = 0.88 | 0.039 (-0.13 to 0.21)  P = 0.65 |
| **Tense feelings** | iSD* | **0.21 (0.01 to 0.42)**  **P = 0.04** | **0.23 (0.03 to 0.44)**  **P = 0.03** |
| **Restless feelings** | iSD* | **0.25 (0.06 to 0.45)**  **P = 0.01** | **0.37 (0.17 to 0.57)**  **P = 0.001** |
| * Mean-adjusted models  The transmasculine group is used as the reference group. | | | |

### 4. Cycle regulation

In Supplementary Table 7, we display differences between the TF group and the TM group on and off cycle regulation at baseline. The results show that the both groups (i.e., TM group with and without cycle regulation) have higher variability in low mood and restless feelings compared to the TF group at baseline.

| **Supplementary Table 7**. Negative affect outcomes before GAHT use (baseline), examining group differences and adjusted for cycle regulation use and psychotropic medication use. The transmasculine group not using cycle regulation is used as the reference group. | | | |
| --- | --- | --- | --- |
|  | | *Transfeminine group vs. transmasculine group - CR* | *Transmasculine + CR vs. transmasculine - CR* |
| **Low mood** | iSD* | **-0.22 (-0.36 to -0.09)**  **P = 0.001** | **-0.20 (-0.36 to -0.04)**  **P = 0.01** |
| **Less interest** | iSD* | -0.04 (-0.17 to 0.10)  P = 0.58 | -0.12 (-0.28 to 0.03)  P = 0.12 |
| **Tense feelings** | iSD* | -0.08 (-0.26 to 0.11)  P = 0.42 | 0.11 (-0.11 to 0.33)  P = 0.31 |
| **Restless feelings** | iSD* | -0.05 (-0.22 to 0.12)  P = 0.53 | 0.09 (-0.10 to 0.28)  P = 0.36 |
| * Mean-adjusted models  CR = cycle regulation | | | |

In Supplementary Table 8, we display the changes in affect variability after correction for cycle regulation in the TM group. The results overall show similar directions and magnitude compared to the results without correction for cycle regulation, although cycle regulation is overall associated with lower variability in low mood.

| **Supplementary Table 8**. Changes in negative affect after GAHT use, stratified per group and adjusted for cycle regulation and psychotropic medication use. The estimated change from baseline and the corresponding 95% confidence intervals and p-values are reported. | | | | |
| --- | --- | --- | --- | --- |
|  | | **Transmasculine group** | | |
|  |  | *3 months of GAHT vs. Baseline* | *12 months of GAHT vs. Baseline* | *CR use vs. no CR use* |
| **Low mood** | iSD* | -0.08 (-0.19 to 0.02)  P = 0.12 | **-0.18 (-0.28 to -0.07)**  **P = 0.002** | **-0.18 (-0.29 to -0.06)**  **P = 0.004** |
| **Less interest** | iSD* | 0.03 (-0.07 to 0.13)  P = 0.53 | 0.03 (-0.07 to 0.14)  P = 0.55 | 0.00 (-0.12 to 0.12)  P = 0.97 |
| **Tense feelings** | iSD* | **-0.17 (-0.31 to -0.03)**  **P = 0.02** | **-0.16 (-0.31 to -0.01)**  **P = 0.04** | 0.01 (-0.14 to 0.17)  P = 0.87 |
| **Restless feelings** | iSD* | -0.10 (-0.26 to-0.05)  P = 0.20 | **-0.22 (-0.38 to -0.06)**  **P = 0.01** | 0.02 (-0.14 to 0.18)  P = 0.82 |
| * Mean-adjusted models | | | | |

### 5. Psychotropic medication

Psychotropic medication was added as covariate to the model – main effects and adjusted group effects at baseline. The longitudinal analyses in the TM group indicate that psychotropic medication use is associated with more variable feelings of less interest. In the longitudinal analyses in the TF group, psychotropic medication use is associated with less variable low mood, less variable tense feelings, and less variable restless feelings. Results are displayed in Supplementary Table 9.

| **Supplementary Table 9**. Negative affect outcomes before GAHT use (baseline), examining group differences and adjusted for cycle regulation use and psychotropic medication use. | | | |
| --- | --- | --- | --- |
|  | | *Transfeminine group vs. transmasculine group* | *Psychotropic medication use vs. no use* |
| **Low mood** | iSD* | **-0.14 (-0.25 to -0.02)**  **P = 0.02** | -0.02 (-0.18 to 0.14)  P = 0.84 |
| **Less interest** | iSD* | 0.02 (-0.10 to 0.13)  P = 0.77 | 0.07 (-0.09 to 0.22)  P = 0.39 |
| **Tense feelings** | iSD* | -0.13 (-0.29 to 0.03)  P = 0.10 | -0.08 (-0.29 to 0.14)  P = 0.48 |
| **Restless feelings** | iSD* | -0.09 (-0.23 to 0.05)  P = 0.20 | 0.06 (-0.14 to 0.25)  P = 0.56 |
| * Mean-adjusted models  The transmasculine group not using cycle regulation is used as the reference group. | | | |

Supplementary Table 10 shows the analyses with the addition of psychotropic medication as a covariate: addition of this covariate does not change the main results.

| **Supplementary Table 10**. Changes in negative affect after GAHT use, stratified per group and adjusted for cycle regulation and psychotropic medication use. | | | | |
| --- | --- | --- | --- | --- |
|  | | **Transmasculine group** | | |
|  |  | *3 months of GAHT vs. baseline* | *12 months of GAHT vs. baseline* | *Psychotropic medication use vs. no use* |
| **Low mood** | iSD* | -0.05 (-0.15 to 0.06)  P = 0.42 | -0.11 (-0.22 to 0.00)  P = 0.05 | 0.12 (-0.05 to 0.29)  P = 0.17 |
| **Less interest** | iSD* | 0.04 (-0.06 to 0.14)  P = 0.45 | 0.04 (-0.07 to 0.14)  P = 0.46 | **0.24 (0.08 to 0.41)**  **P = 0.01** |
| **Tense feelings** | iSD* | **-0.18 (-0.32 to -0.04)**  **P = 0.01** | **-0.18 (-0.32 to -0.04)**  **P = 0.02** | 0.04 (-0.17 to 0.24)  P = 0.73 |
| **Restless feelings** | iSD* | -0.078 (-0.23 to 0.07)  P = 0.32 | **-0.21 (-0.36 to -0.06)**  **P = 0.01** | 0.15 (-0.04 to 0.33)  P = 0.12 |
|  | | **Transfeminine group** | | |
|  |  | *3 months of GAHT vs. baseline* | *12 months of GAHT vs. baseline* | *Psychotropic medication use vs. no use* |
| **Low mood** | iSD* | **0.19 (0.07 to 0.32)**  **P = 0.004** | **0.13 (0.00 to 0.26)**  **P = 0.06** | **-0.20 (-0.40 to 0.00)**  **P = 0.05** |
| **Less interest** | iSD* | 0.06 (-0.07 to 0.19)  P = 0.38 | 0.07 (-0.06 to 0.20)  P = 0.31 | 0.14 (-0.34 to 0.07)  P = 0.20 |
| **Tense feelings** | iSD* | 0.05 (-0.10 to 0.19)  P = 0.55 | 0.06 (-0.09 to 0.21)  P = 0.41 | **-0.27 (-0.47 to -0.08)**  **P = 0.01** |
| **Restless feelings** | iSD* | **0.13 (0.00 to 0.26)**  **P = 0.05** | 0.05 (-0.09 to 0.18)  P = 0.49 | **-0.33 (-0.52 to -0.14)**  **P = 0.001** |
| * Mean-adjusted models  All cells display the mean and SD of each iMean and iSD, and in outcomes which show significant changes, the estimated change from baseline and the corresponding 95% confidence intervals and p-values are reported. Red represents decreases and green represents increases. | | | | |
